# Comorbidities, clinical signs and symptoms, laboratory findings, imaging features, treatment strategies, and outcomes in adult and pediatric patients with COVID-19: A systematic review and meta-analysis

**DOI:** 10.1101/2020.05.20.20103804

**Authors:** Catherine R. Jutzeler, Lucie Bourguignon, Caroline V. Weis, Bobo Tong, Cyrus Wong, Bastian Rieck, Hans Pargger, Sarah Tschudin-Sutter, Adrin Egli, Karsten Borgwardt, Matthias Walter

**Affiliations:** Department of Biosystems Science and Engineering, ETH Zurich, Basel, Switzerland; SIB Swiss Institute of Bioinformatics, Lausanne, Switzerland; Spinal Cord Injury Center, Balgrist University Hospital, University of Zurich, Zurich, Switzerland; International Collaboration on Repair Discoveries (ICOR, University of British Columbia, Vancouver, Canada; Simon Fraser University, Vancouver, Canada; Intensive Care Unit, University Hospital Basel, University Basel, Basel, Switzerland; Division of Infectious Diseases & Hospital Epidemiology, University Hospital Basel and University of Basel, Switzerland; Department of Clinical Research, University Hospital Basel and University of Basel, Switzerland; Division of Clinical Bacteriology & Mycology, University Hospital Basel, Basel, Switzerland; Applied Microbiology Research, Department of Biomedicine, University of Basel, Basel, Switzerland; Swiss Paraplegic Center, Nottwil, Switzerland

**Keywords:** SARS-CoV-2, COVID-19, meta-analysis, systematic review, comorbidities, clinical characteristics, laboratory findings, imaging features, treatment

## Abstract

**Introduction:** Since December 2019, a novel coronavirus (SARS-CoV-2) has triggered a world-wide pandemic with an enormous medical, societal, and economic toll. Thus, our aim was to gather all available information regarding comorbidities, clinical signs and symptoms, outcomes, laboratory findings, imaging features, and treatments in patients with coronavirus disease 2019 (COVID-19).

**Methods:** EMBASE, PubMed/ Medline, Scopus, and Web of Science were searched for studies published in any language between December 1st, 2019 and March 28th. Original studies were included if the exposure of interest was an infection with SARS-CoV-2 or confirmed COVID-19. The primary outcome was the risk ratio of comorbidities, clinical signs and symptoms, imaging features, treatments, outcomes, and complications associated with COVID-19 morbidity and mortality. We performed random-effects pairwise meta-analyses for proportions and relative risks, I2, Tau2, and Cochrane Q, sensitivity analyses, and assessed publication bias.

**Results:** 148 met the inclusion criteria for the systematic review and meta-analysis with 12’149 patients (5’739 female) and a median age was 47.0 [35.0–64.6]. 617 patients died from COVID-19 and its complication, while 297 patients were reported as asymptomatic. Older age (SMD: 1.25 [0.78-1.72]; p < 0.001), being male (RR = 1.32 [1.13–1.54], p = 0.005) and pre-existing comorbidity (RR = 1.69 [1.48–1.94]; p < 0.001) were identified as risk factors of in-hospital mortality. The heterogeneity between studies varied substantially (I2; range: 1.5–98.2%). Publication bias was only found in eight studies (Egger’s test: p < 0.05).

**Conclusions:** Our meta-analyses revealed important risk factors that are associated with severity and mortality of COVID-19.

## 1. Introduction

The severe acute respiratory syndrome (SARS) coronavirus 2 (SARS-CoV-2) initially emerged in Wuhan, Hubei, People’s Republic of China and has been identified as the causative agent of coronavirus disease 2019 (COVID-19). It’s pandemic spread presents a substantial medical challenge with an enormous societal and economic toll^1,2^. Similar to influenza and SARS-CoV-1, SARS-CoV-2 is considered a “crowd disease” that spreads most easily when individuals are packed together at high densities. Phylogenetic data implicate a zoonotic origin^3^ and the rapid spread suggests ongoing person-to-person transmission^4^. Additional factors contributing to the rapid spread constitute the duration of the incubation period^5^ and infectiousness peaking on or before symptom onset^6^ contribute to the rapid spread of SARS-CoV-2. Another factor contributing to the rapid spread and alarmingly high number of infected people is the SARS-CoV-2 nature of initial dormancy of symptoms. The most common symptoms associated with COVID-19 include a sudden onset of fever, coughing, and dyspnea^2,7,8^. Complications comprise acute respiratory distress syndrome (ARDS), pneumonia, kidney failure, bacterial superinfections, coagulation abnormalities and thromboembolic events, sepsis, and even death^9,10^. So far, only a few demographic and clinical factors, such as older age, diabetes, and cardiovascular diseases, have been linked with poor outcome and increased risk of mortality^11,12^. This knowledge gap extends to the risk of infections, disease progression, and outcome in vulnerable patient populations, including newborns, children, pregnant, and elderly patients. A better understanding of the risks for these vulnerable patient populations is critical in order to optimize their protection and tailor prevention and treatment strategies. Thus, the aim of our systematic review and meta-analysis was to gather available information in the literature and determine the most prevalent comorbidities, clinical signs and symptoms, imaging features, laboratory parameters, treatments, outcomes, and complications arising in patients with COVID-19. We stratified our systematic reviews and meta-analysis by different cohorts, namely pediatric/neonatal and adult COVID-19 patients including pregnant women. Furthermore, we aimed to assess current evidence for the associations between risk factors and in-hospital mortality. Based on previous reports, we addressed the hypothesis that male sex, older age, as well as pre-existing hypertension and diabetes mellitus are risk factors of morbidity and mortality in patients with COVID-19.

## 2. Methods

Our systematic review and meta-analysis adhere to the Preferred Reporting Items for Systematic reviews and Meta-Analysis (PRISMA) statement^13^ and Meta-analysis of Observational Studies in Epidemiology (MOOSE) checklist^14^.

### 2.1. Search strategy and selection criteria

Four bibliographic databases were systematically searched: EMBASE, PubMed/ Medline, Scopus, and Web of Science. Our search was not restricted by language. We searched for studies published from December 1st, 2019 to March 28^th^, 2020, with search terms related to COVID-19 (“COVID-19”, “SARS-CoV-2”, “coronavirus disease 2019”, “severe acute respiratory syndrome coronavirus 2”, “2019 novel coronavirus”, “2019-nCoV”, “coronavirus”, and “corona virus”). The full search strategy is provided in **Appendix 1.** Manual searching was also performed, reviewing reference lists of relevant studies and comprehensive review articles. Records were managed by EndNote X 8.0 software to exclude duplicates.

### 2.2. Selection of studies

Two investigators (CRJ and MW) independently screened the titles and abstracts to determine whether studies should be included. Eligibility criteria were also applied to the full-text articles during the final selection. In case multiple articles reported on a single study, the article that provided the most data was selected for further synthesis. We quantified the inter-rater agreement for study selection using Cohen’s κ coefficient^15^. Articles written in Chinese were reviewed by our two native speaking authors (BT and CW) and if the inclusion criteria were met, these authors also extracted the specified data. All disagreements were discussed and resolved at a consensus meeting.

### 2.3. Inclusion and exclusion criteria

All full-text, peer-reviewed articles that described case-control, cohort studies, or case studies investigating the epidemiological and clinical features, comorbidities, laboratory parameters, imaging features, and/or treatment of patients that were diagnosed with COVID-19. We excluded duplicate publications, non-peer reviewed articles (e.g., preprints), reviews, meta-analyses, abstracts or conference proceedings, editorials, commentaries, letters with insufficient data, studies on non-human species, or out-of-scope studies (e.g., comparison with other infections, case-fatality reports). In case multiple studies published data from the same cohort, we included the article representing the most inclusive information on the population to avoid overlap. Lastly, studies that did not report demographics (i.e., age and sex) were also excluded. Figure 1 outlines our search strategy and application of inclusion and exclusion criteria.

**Figure 1.**
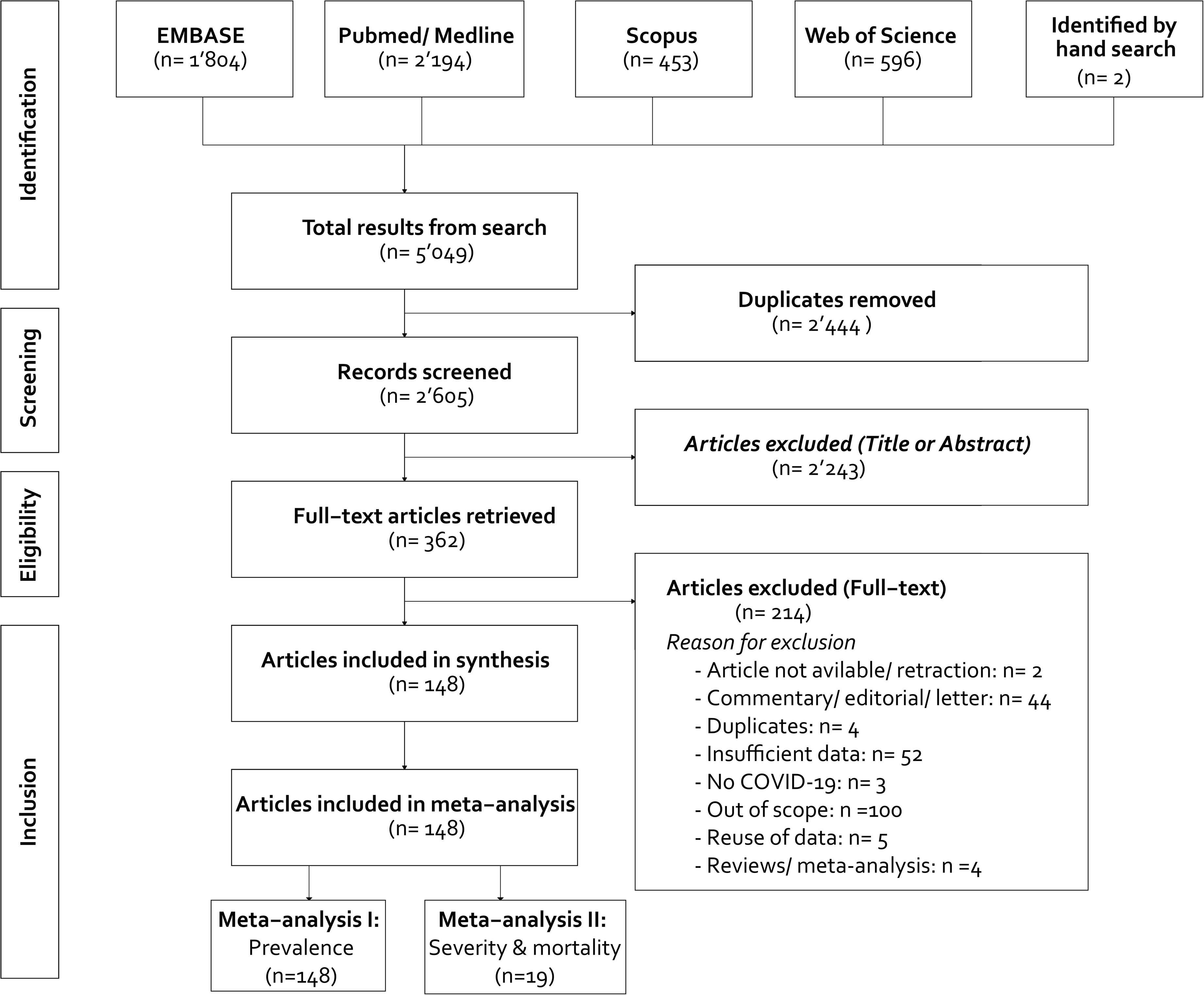
Flow-chart of the search strategy. A total of 148 studies were eligible for the literature review and the first part of the meta-analysis (i.e., prevalence). Nine-teen studies were included in the second part of the meta-analysis (i.e., severity and mortality).

### 2.4. Data extraction and synthesis

Data extraction tables were created with the following information: 1) publication information (i.e., author, date, language of article, country where the study was performed, study design [case study, case series, or cohort study]^16^, study population [pediatric/neonatal and adult COVID-19 patients including pregnant women); 2) demographics (i.e., age, sex); 3) clinical signs and symptoms (e.g., cough, fatigue, fever, sputum); 3) comorbidities (e.g., hypertension, diabetes, cardiovascular diseases); 4) therapies administered to treat COVID-19 (e.g., antibiotics, antivirals, invasive mechanical ventilation); 5) clinical outcomes (e.g., death, survival, recovery); and 6) complications associated with COVID-19 (e.g., sepsis and shock, ARDS). In case studies provided data for multiple patient groups(e.g., pediatric and adult patient), we extracted this information separately for each group. A full list of extracted variables is provided in Supplementary Table 1.

**Table 1.**
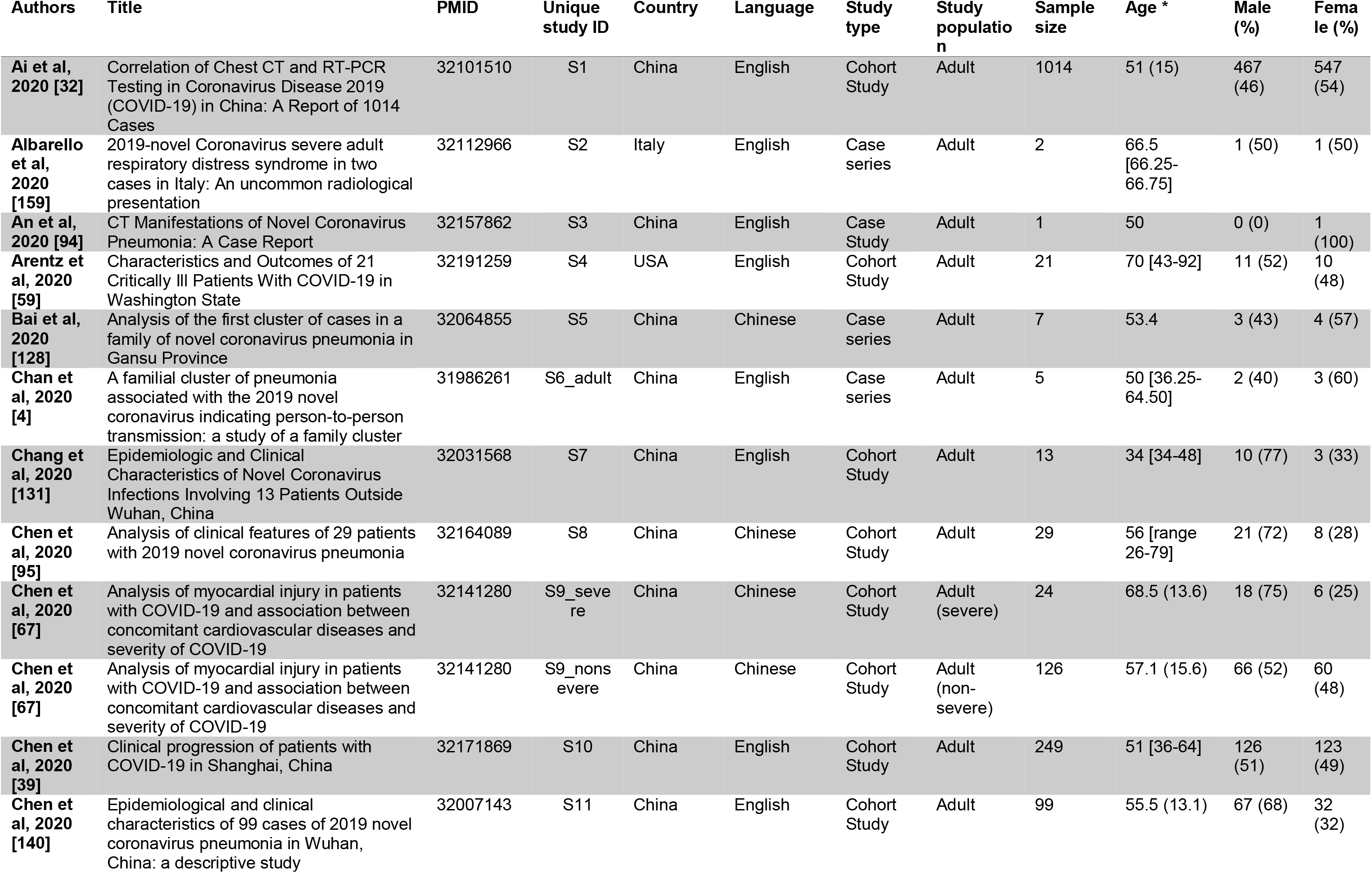
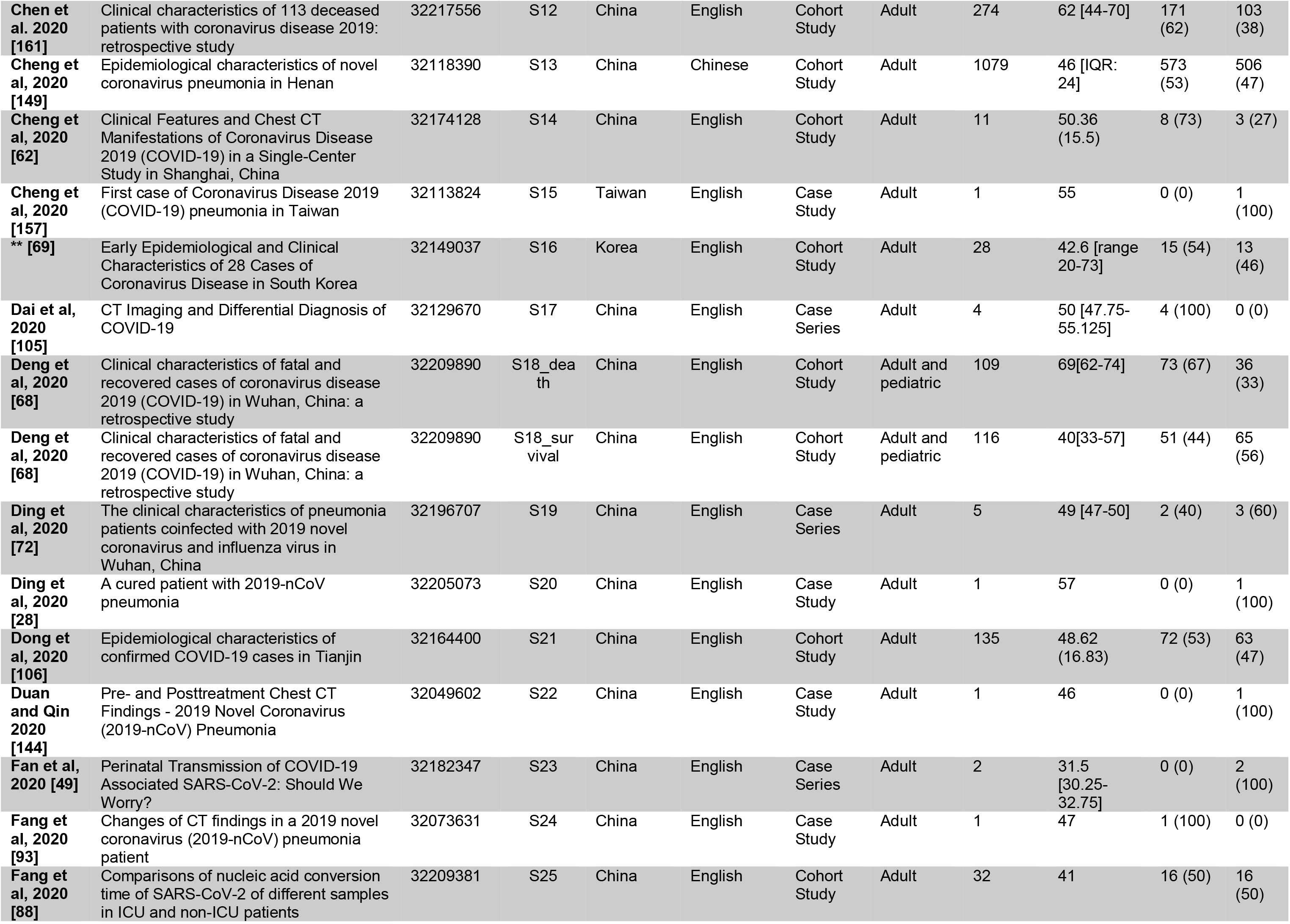
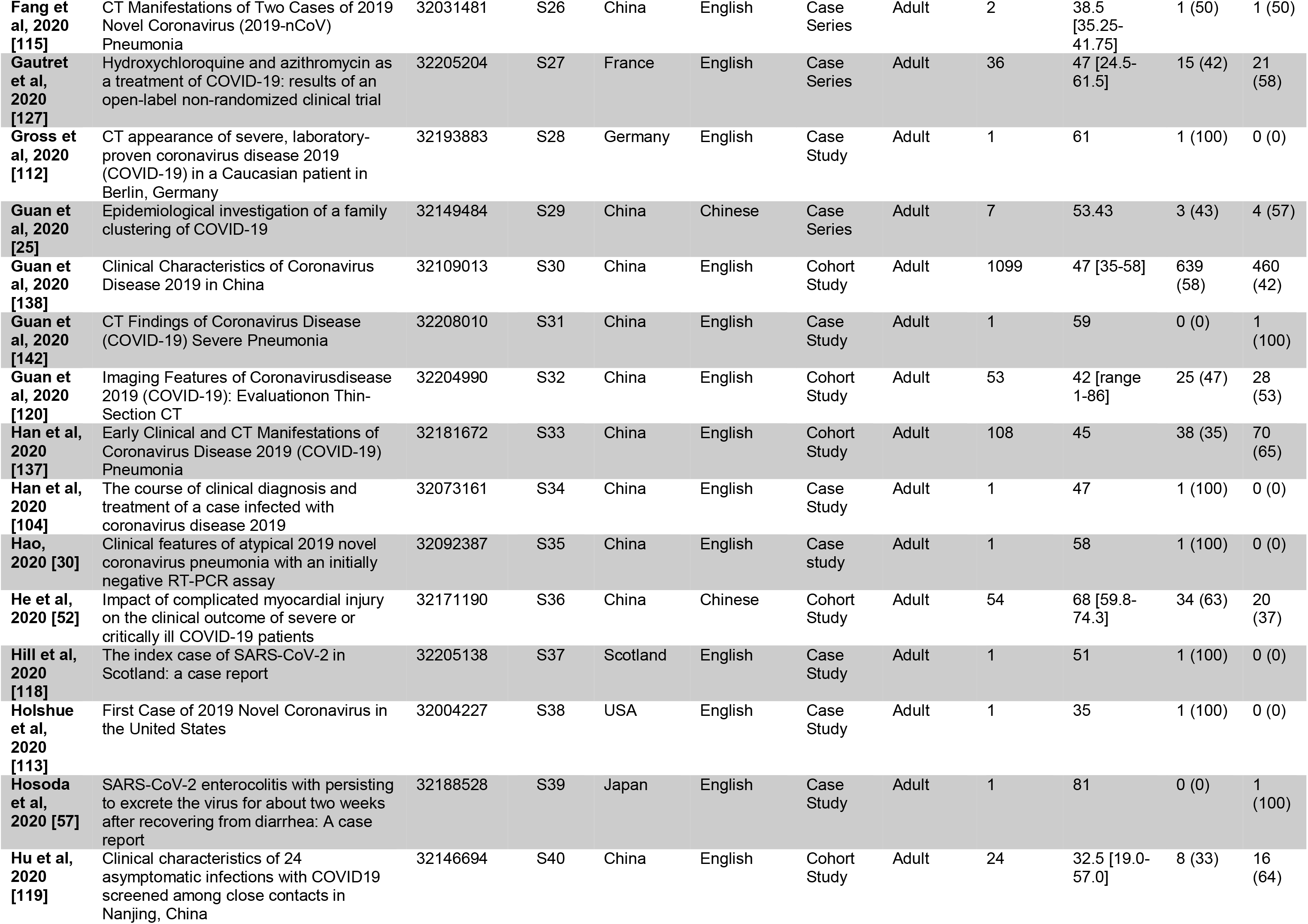
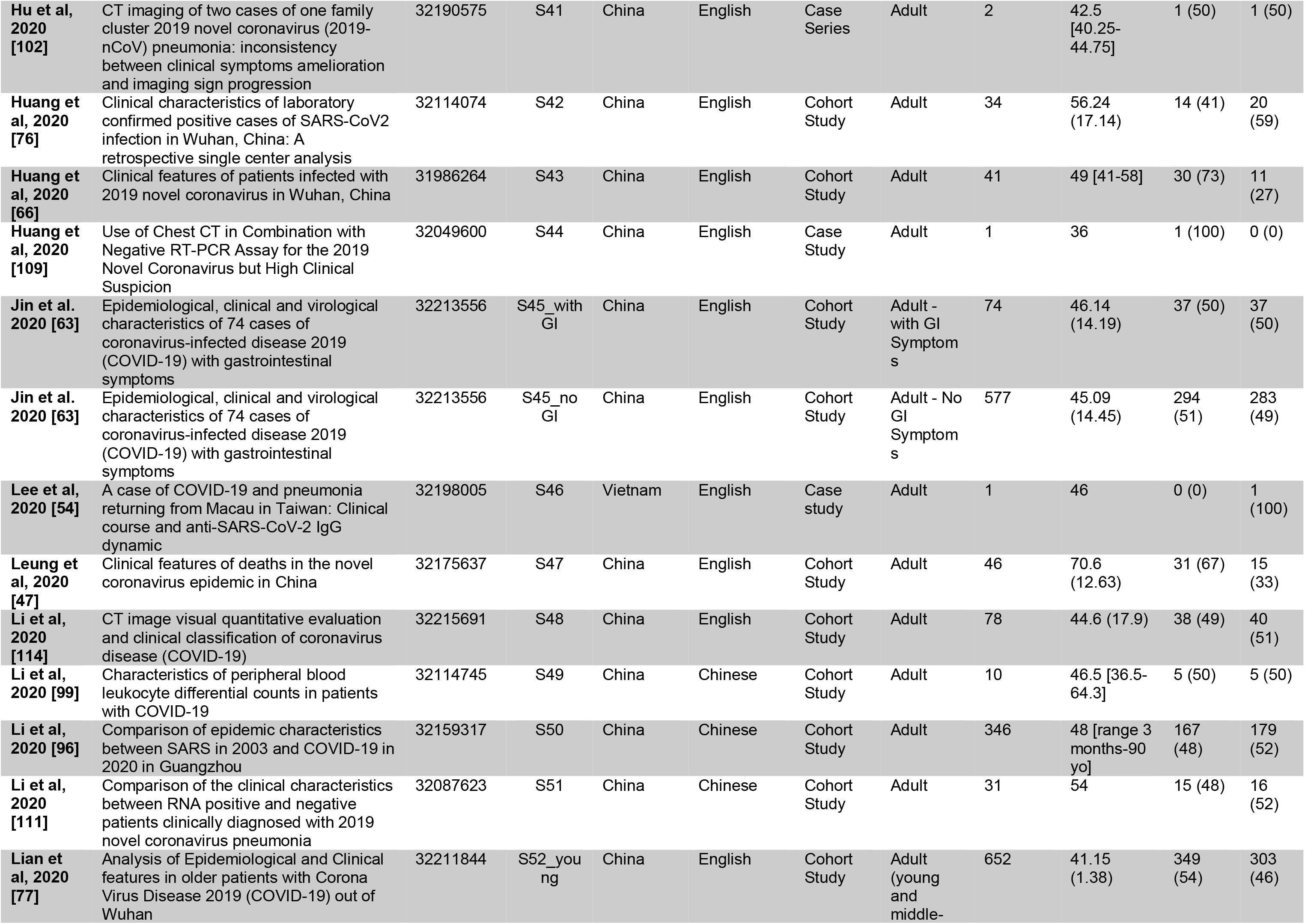
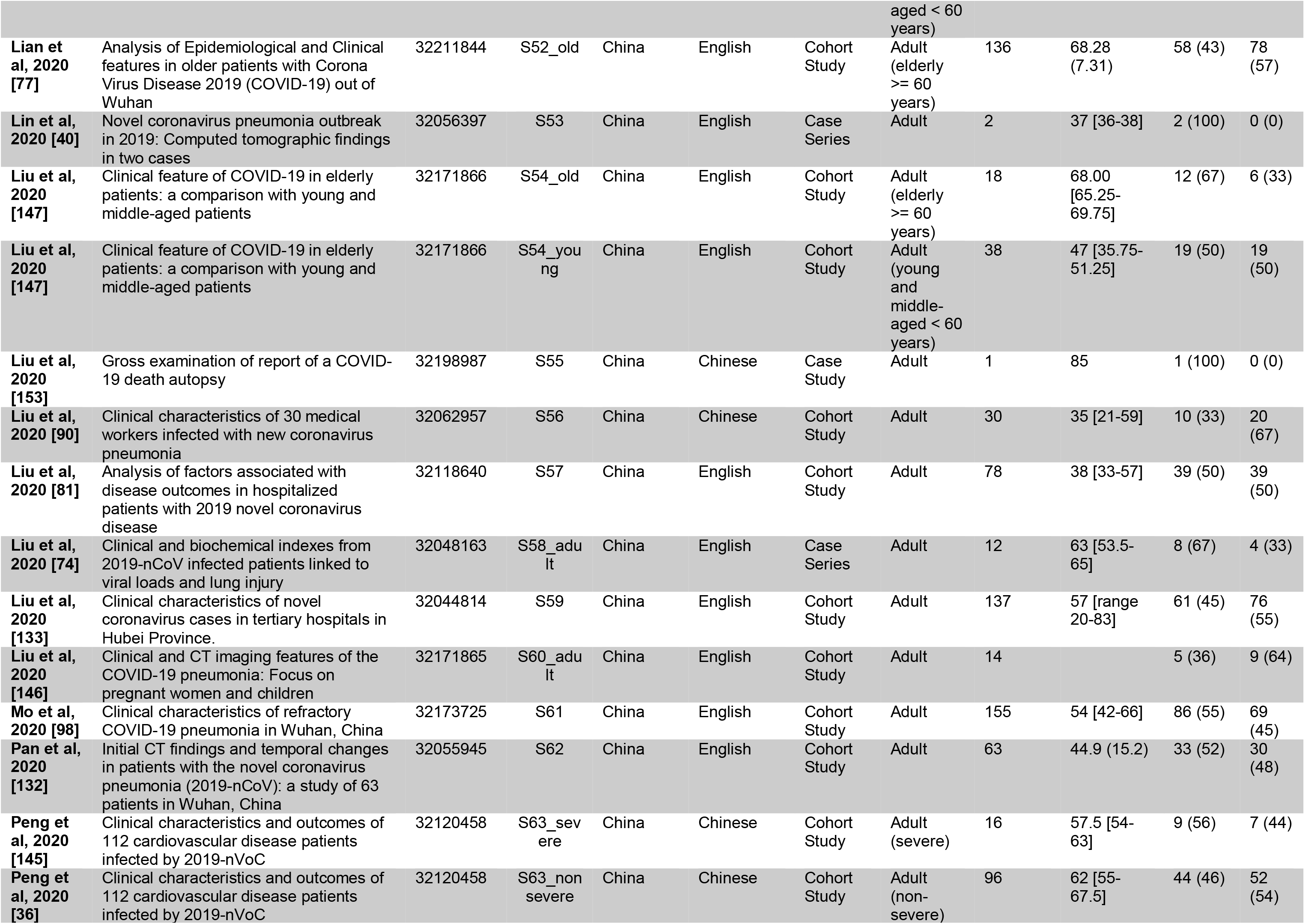
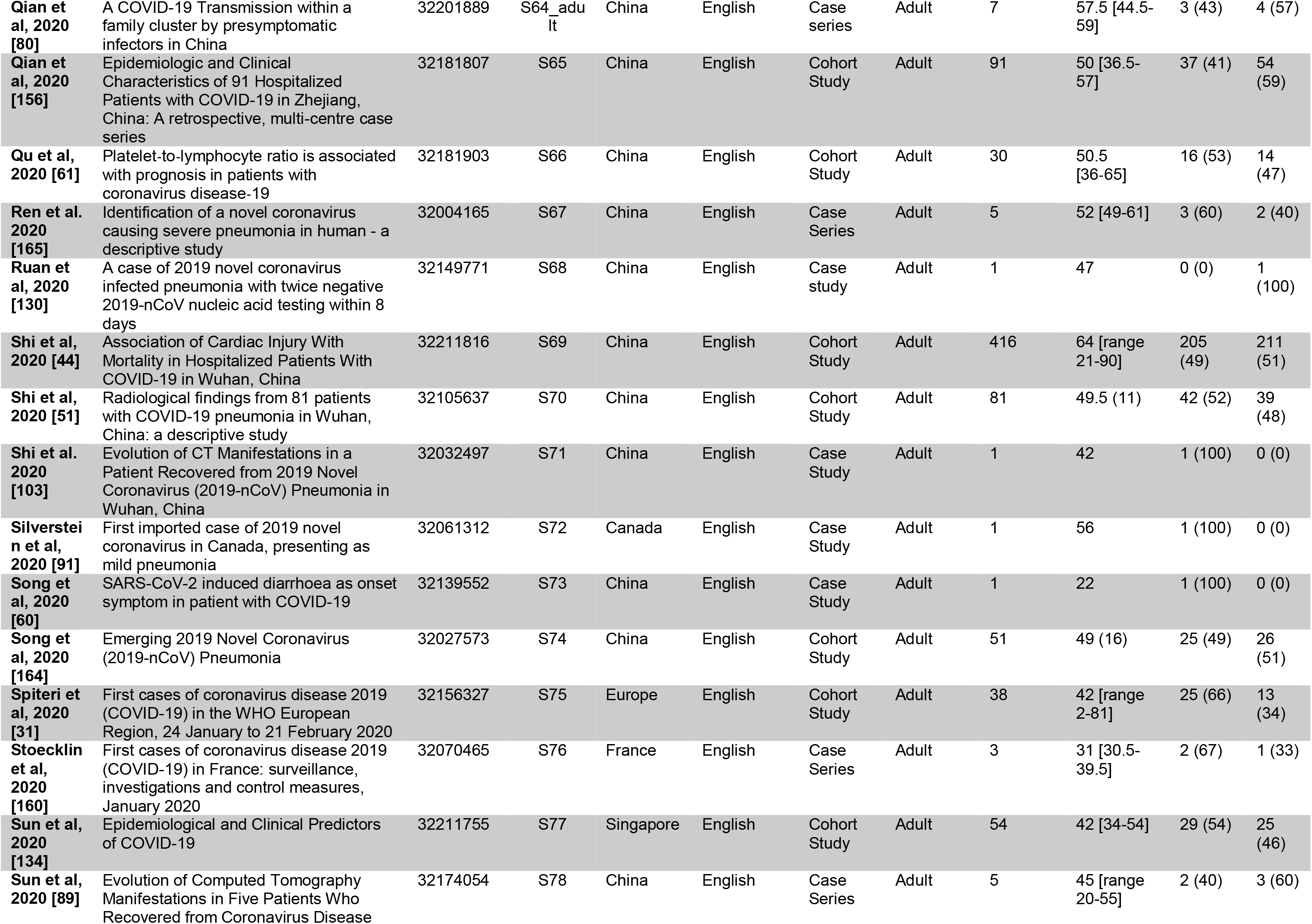
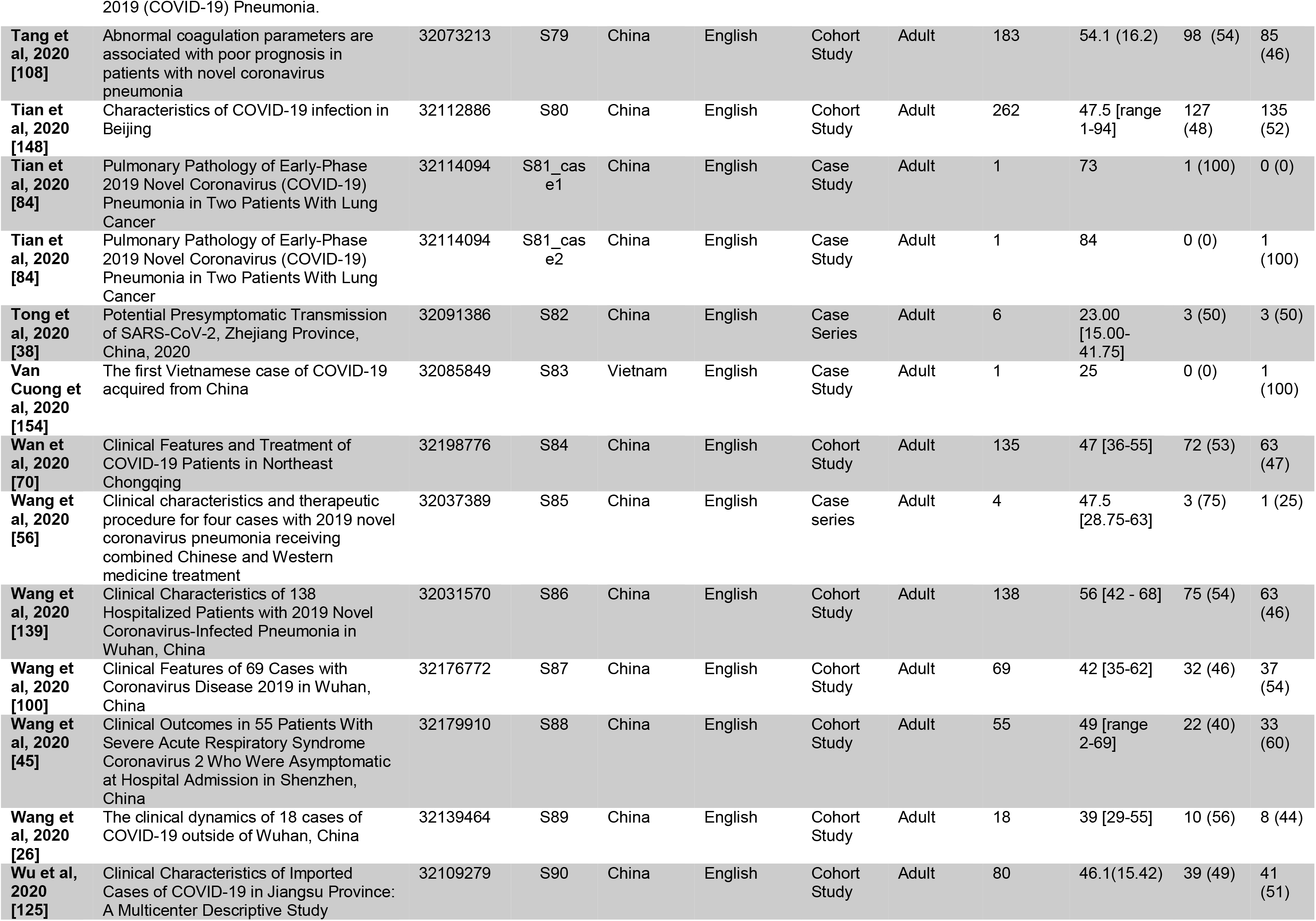
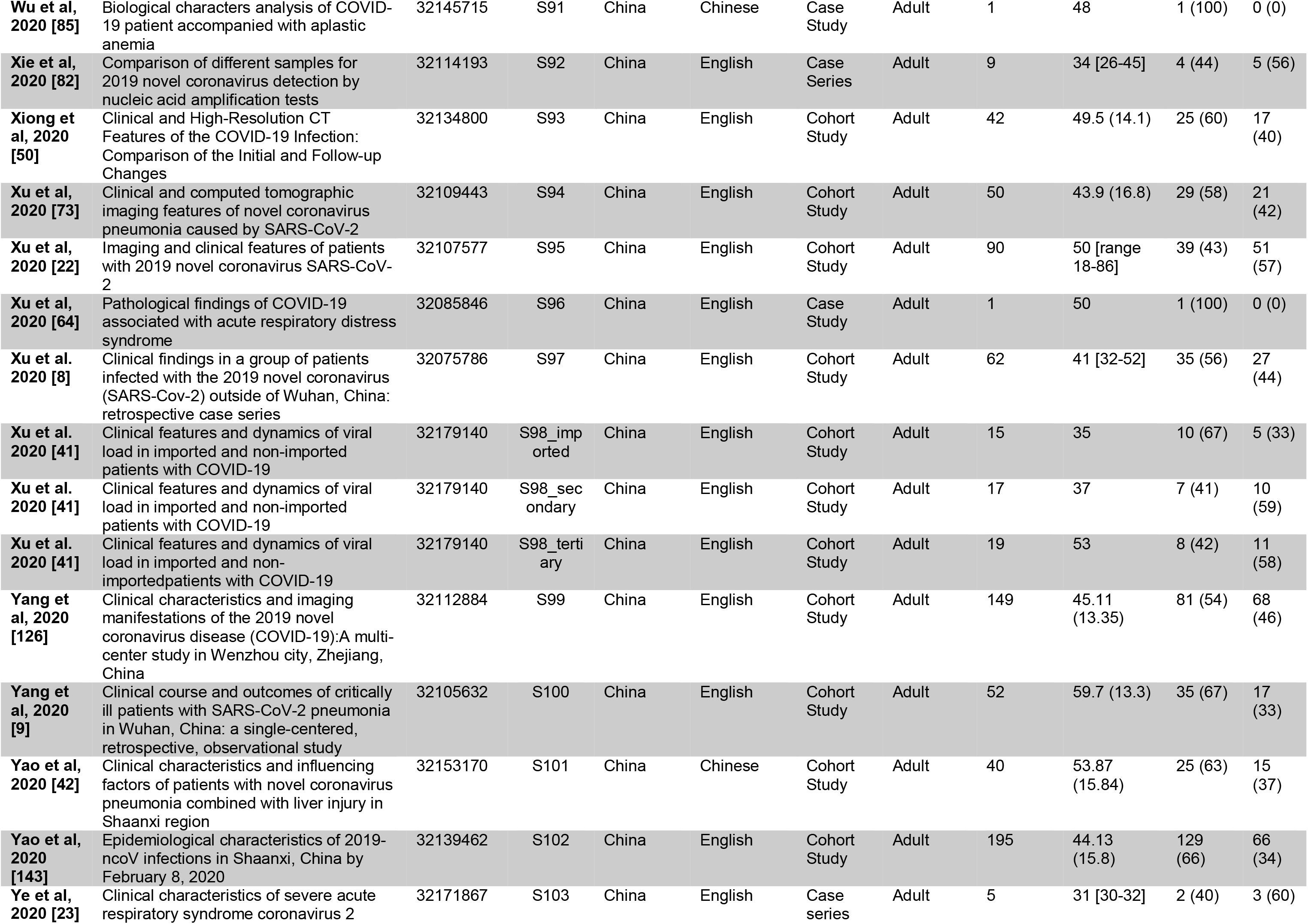
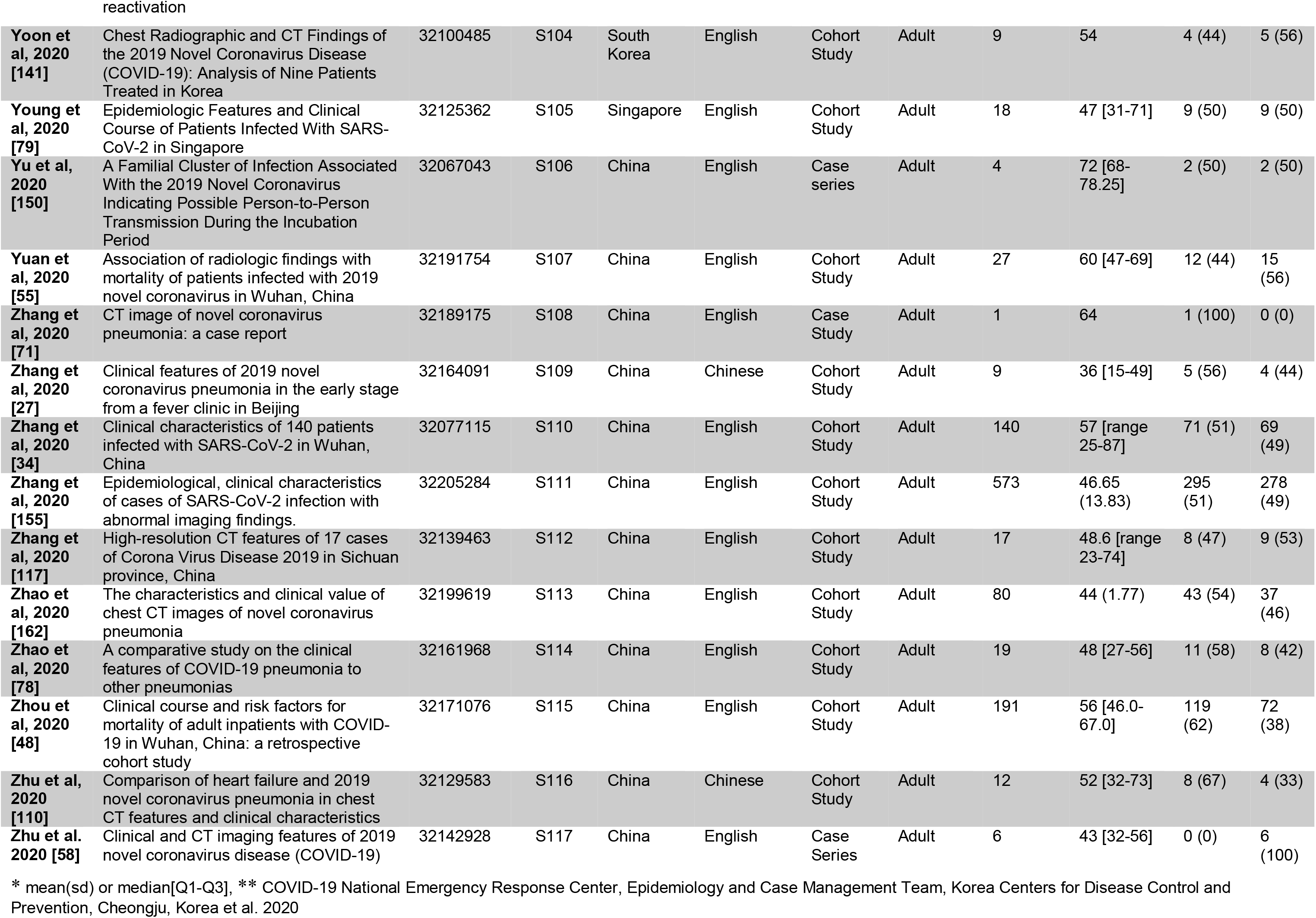
Included studies of adults with COVID-19.

| Authors | Title | PMID | Unique study ID | Country | Language | Study type | Study population | Sample size | Age * | Male (%) | Female (%) |
| --- | --- | --- | --- | --- | --- | --- | --- | --- | --- | --- | --- |
| **Ai et al, 2020 [32]** | Correlation of Chest CT and RT-PCR Testing in Coronavirus Disease 2019 (COVID-19) in China: A Report of 1014 Cases | 32101510 | S1 | China | English | Cohort Study | Adult | 1014 | 51 (15) | 467 (46) | 547 (54) |
| **Albareello et al, 2020 [159]** | 2019-novel Coronavirus severe adult respiratory distress syndrome in two cases in Italy: An uncommon radiological presentation | 32112966 | S2 | Italy | English | Case series | Adult | 2 | 66.5 [66.25-66.75] | 1 (50) | 1 (50) |
| **An et al, 2020 [94]** | CT Manifestations of Novel Coronavirus Pneumonia: A Case Report | 32157862 | S3 | China | English | Case Study | Adult | 1 | 50 | 0 (0) | 1 (100) |
| **Arentz et al, 2020 [59]** | Characteristics and Outcomes of 21 Critically Ill Patients With COVID-19 in Washington State | 32191259 | S4 | USA | English | Cohort Study | Adult | 21 | 70 [43-92] | 11 (52) | 10 (48) |
| **Bai et al, 2020 [128]** | Analysis of the first cluster of cases in a family of novel coronavirus pneumonia in Gansu Province | 32064855 | S5 | China | Chinese | Case series | Adult | 7 | 53.4 | 3 (43) | 4 (57) |
| **Chan et al, 2020 [4]** | A familial cluster of pneumonia associated with the 2019 novel coronavirus indicating person-to-person transmission: a study of a family cluster | 31986261 | S6_adult | China | English | Case series | Adult | 5 | 50 [36.25-64.50] | 2 (40) | 3 (60) |
| **Chang et al, 2020 [131]** | Epidemiologic and Clinical Characteristics of Novel Coronavirus Infections Involving 13 Patients Outside Wuhan, China | 32031568 | S7 | China | English | Cohort Study | Adult | 13 | 34 [34-48] | 10 (77) | 3 (33) |
| **Chen et al, 2020 [95]** | Analysis of clinical features of 29 patients with 2019 novel coronavirus pneumonia | 32164089 | S8 | China | Chinese | Cohort Study | Adult | 29 | 56 [range 26-79] | 21 (72) | 8 (28) |
| **Chen et al, 2020 [67]** | Analysis of myocardial injury in patients with COVID-19 and association between concomitant cardiovascular diseases and severity of COVID-19 | 32141280 | S9_severe | China | Chinese | Cohort Study | Adult (severe) | 24 | 68.5 (13.6) | 18 (75) | 6 (25) |
| **Chen et al, 2020 [67]** | Analysis of myocardial injury in patients with COVID-19 and association between concomitant cardiovascular diseases and severity of COVID-19 | 32141280 | S9_nonsvere | China | Chinese | Cohort Study | Adult (non-severe) | 126 | 57.1 (15.6) | 66 (52) | 60 (48) |
| **Chen et al, 2020 [39]** | Clinical progression of patients with COVID-19 in Shanghai, China | 32171869 | S10 | China | English | Cohort Study | Adult | 249 | 51 [36-64] | 126 (51) | 123 (49) |
| **Chen et al, 2020 [140]** | Epidemiological and clinical characteristics of 99 cases of 2019 novel coronavirus pneumonia in Wuhan, China: a descriptive study | 32007143 | S11 | China | English | Cohort Study | Adult | 99 | 55.5 (13.1) | 67 (68) | 32 (32) |
| **Chen et al, 2020 [161]** | Clinical characteristics of 113 deceased patients with coronavirus disease 2019: retrospective study | 32217556 | S12 | China | English | Cohort Study | Adult | 274 | 62 [44-70] | 171 (62) | 103 (38) |
| **Cheng et al, 2020 [149]** | Epidemiological characteristics of novel coronavirus pneumonia in Henan | 32118390 | S13 | China | Chinese | Cohort Study | Adult | 1079 | 46 [IQR: 24] | 573 (53) | 506 (47) |
| **Cheng et al, 2020 [62]** | Clinical Features and Chest CT Manifestations of Coronavirus Disease 2019 (COVID-19) in a Single-Center Study in Shanghai, China | 32174128 | S14 | China | English | Cohort Study | Adult | 11 | 50.36 (15.5) | 8 (73) | 3 (27) |
| **Cheng et al, 2020 [157]** | First case of Coronavirus Disease 2019 (COVID-19) pneumonia in Taiwan | 32113824 | S15 | Taiwan | English | Case Study | Adult | 1 | 55 | 0 (0) | 1 (100) |
| **\*\* [69]** | Early Epidemiological and Clinical Characteristics of 28 Cases of Coronavirus Disease in South Korea | 32149037 | S16 | Korea | English | Cohort Study | Adult | 28 | 42.6 [range 20-73] | 15 (54) | 13 (46) |
| **Dai et al, 2020 [105]** | CT Imaging and Differential Diagnosis of COVID-19 | 32129670 | S17 | China | English | Case Series | Adult | 4 | 50 [47.75-55.125] | 4 (100) | 0 (0) |
| **Deng et al, 2020 [68]** | Clinical characteristics of fatal and recovered cases of coronavirus disease 2019 (COVID-19) in Wuhan, China: a retrospective study | 32209890 | S18_death | China | English | Cohort Study | Adult and pediatric | 109 | 69[62-74] | 73 (67) | 36 (33) |
| **Deng et al, 2020 [68]** | Clinical characteristics of fatal and recovered cases of coronavirus disease 2019 (COVID-19) in Wuhan, China: a retrospective study | 32209890 | S18_survival | China | English | Cohort Study | Adult and pediatric | 116 | 40[33-57] | 51 (44) | 65 (56) |
| **Ding et al, 2020 [72]** | The clinical characteristics of pneumonia patients coinfectd with 2019 novel coronavirus and influenza virus in Wuhan, China | 32196707 | S19 | China | English | Case Series | Adult | 5 | 49 [47-50] | 2 (40) | 3 (60) |
| **Ding et al, 2020 [28]** | A cured patient with 2019-nCoV pneumonia | 32205073 | S20 | China | English | Case Study | Adult | 1 | 57 | 0 (0) | 1 (100) |
| **Dong et al, 2020 [106]** | Epidemiological characteristics of confirmed COVID-19 cases in Tianjin | 32164400 | S21 | China | English | Cohort Study | Adult | 135 | 48.62 (16.83) | 72 (53) | 63 (47) |
| **Duan and Qin 2020 [144]** | Pre- and Posttreatment Chest CT Findings - 2019 Novel Coronavirus (2019-nCoV) Pneumonia | 32049602 | S22 | China | English | Case Study | Adult | 1 | 46 | 0 (0) | 1 (100) |
| **Fan et al, 2020 [49]** | Perinatal Transmission of COVID-19 Associated SARS-CoV-2: Should We Worry? | 32182347 | S23 | China | English | Case Series | Adult | 2 | 31.5 [30.25-32.75] | 0 (0) | 2 (100) |
| **Fang et al, 2020 [93]** | Changes of CT findings in a 2019 novel coronavirus (2019-nCoV) pneumonia patient | 32073631 | S24 | China | English | Case Study | Adult | 1 | 47 | 1 (100) | 0 (0) |
| **Fang et al, 2020 [88]** | Comparisons of nucleic acid conversion time of SARS-CoV-2 of different samples in ICU and non-ICU patients | 32209381 | S25 | China | English | Cohort Study | Adult | 32 | 41 | 16 (50) | 16 (50) |
| **Fang et al, 2020 [115]** | CT Manifestations of Two Cases of 2019 Novel Coronavirus (2019-nCoV) Pneumonia | 32031481 | S26 | China | English | Case Series | Adult | 2 | 38.5 [35.25-41.75] | 1 (50) | 1 (50) |
| **Gautret et al, 2020 [127]** | Hydroxychloroquine and azithromycin as a treatment of COVID-19: results of an open-label non-randomized clinical trial | 32205204 | S27 | France | English | Case Series | Adult | 36 | 47 [24.5-61.5] | 15 (42) | 21 (58) |
| **Gross et al, 2020 [112]** | CT appearance of severe, laboratory-proven coronavirus disease 2019 (COVID-19) in a Caucasian patient in Berlin, Germany | 32193883 | S28 | Germany | English | Case Study | Adult | 1 | 61 | 1 (100) | 0 (0) |
| **Guan et al, 2020 [25]** | Epidemiological investigation of a family clustering of COVID-19 | 32149484 | S29 | China | Chinese | Case Series | Adult | 7 | 53.43 | 3 (43) | 4 (57) |
| **Guan et al, 2020 [138]** | Clinical Characteristics of Coronavirus Disease 2019 in China | 32109013 | S30 | China | English | Cohort Study | Adult | 1099 | 47 [35-58] | 639 (58) | 460 (42) |
| **Guan et al, 2020 [142]** | CT Findings of Coronavirus Disease (COVID-19) Severe Pneumonia | 32208010 | S31 | China | English | Case Study | Adult | 1 | 59 | 0 (0) | 1 (100) |
| **Guan et al, 2020 [120]** | Imaging Features of Coronavirusdisease 2019 (COVID-19): Evaluationon Thin-Section CT | 32204990 | S32 | China | English | Cohort Study | Adult | 53 | 42 [range 1-86] | 25 (47) | 28 (53) |
| **Han et al, 2020 [137]** | Early Clinical and CT Manifestations of Coronavirus Disease 2019 (COVID-19) Pneumonia | 32181672 | S33 | China | English | Cohort Study | Adult | 108 | 45 | 38 (35) | 70 (65) |
| **Han et al, 2020 [104]** | The course of clinical diagnosis and treatment of a case infected with coronavirus disease 2019 | 32073161 | S34 | China | English | Case Study | Adult | 1 | 47 | 1 (100) | 0 (0) |
| **Hao, 2020 [30]** | Clinical features of atypical 2019 novel coronavirus pneumonia with an initially negative RT-PCR assay | 32092387 | S35 | China | English | Case study | Adult | 1 | 58 | 1 (100) | 0 (0) |
| **He et al, 2020 [52]** | Impact of complicated myocardial injury on the clinical outcome of severe or critically ill COVID-19 patients | 32171190 | S36 | China | Chinese | Cohort Study | Adult | 54 | 68 [59.8-74.3] | 34 (63) | 20 (37) |
| **Hill et al, 2020 [118]** | The index case of SARS-CoV-2 in Scotland: a case report | 32205138 | S37 | Scotland | English | Case Study | Adult | 1 | 51 | 1 (100) | 0 (0) |
| **Holshue et al, 2020 [113]** | First Case of 2019 Novel Coronavirus in the United States | 32004227 | S38 | USA | English | Case Study | Adult | 1 | 35 | 1 (100) | 0 (0) |
| **Hosoda et al, 2020 [57]** | SARS-CoV-2 enterocolitis with persisting to excrete the virus for about two weeks after recovering from diarrhea: A case report | 32188528 | S39 | Japan | English | Case Study | Adult | 1 | 81 | 0 (0) | 1 (100) |
| **Hu et al, 2020 [119]** | Clinical characteristics of 24 asymptomatic infections with COVID19 screened among close contacts in Nanjing, China | 32146694 | S40 | China | English | Cohort Study | Adult | 24 | 32.5 [19.0-57.0] | 8 (33) | 16 (64) |
| **Hu et al, 2020 [102]** | CT imaging of two cases of one family cluster 2019 novel coronavirus (2019-nCoV) pneumonia: inconsistency between clinical symptoms amelioration and imaging sign progression | 32190575 | S41 | China | English | Case Series | Adult | 2 | 42.5 [40.25-44.75] | 1 (50) | 1 (50) |
| **Huang et al, 2020 [76]** | Clinical characteristics of laboratory confirmed positive cases of SARS-CoV2 infection in Wuhan, China: A retrospective single center analysis | 32114074 | S42 | China | English | Cohort Study | Adult | 34 | 56.24 (17.14) | 14 (41) | 20 (59) |
| **Huang et al, 2020 [66]** | Clinical features of patients infected with 2019 novel coronavirus in Wuhan, China | 31986264 | S43 | China | English | Cohort Study | Adult | 41 | 49 [41-58] | 30 (73) | 11 (27) |
| **Huang et al, 2020 [109]** | Use of Chest CT in Combination with Negative RT-PCR Assay for the 2019 Novel Coronavirus but High Clinical Suspicion | 32049600 | S44 | China | English | Case Study | Adult | 1 | 36 | 1 (100) | 0 (0) |
| **Jin et al. 2020 [63]** | Epidemiological, clinical and virological characteristics of 74 cases of coronavirus-infected disease 2019 (COVID-19) with gastrointestinal symptoms | 32213556 | S45_with GI | China | English | Cohort Study | Adult - with GI Symptoms | 74 | 46.14 (14.19) | 37 (50) | 37 (50) |
| **Jin et al. 2020 [63]** | Epidemiological, clinical and virological characteristics of 74 cases of coronavirus-infected disease 2019 (COVID-19) with gastrointestinal symptoms | 32213556 | S45_no GI | China | English | Cohort Study | Adult - No GI Symptoms | 577 | 45.09 (14.45) | 294 (51) | 283 (49) |
| **Lee et al, 2020 [54]** | A case of COVID-19 and pneumonia returning from Macau in Taiwan: Clinical course and anti-SARS-CoV-2 IgG dynamic | 32198005 | S46 | Vietnam | English | Case study | Adult | 1 | 46 | 0 (0) | 1 (100) |
| **Leung et al, 2020 [47]** | Clinical features of deaths in the novel coronavirus epidemic in China | 32175637 | S47 | China | English | Cohort Study | Adult | 46 | 70.6 (12.63) | 31 (67) | 15 (33) |
| **Li et al, 2020 [114]** | CT image visual quantitative evaluation and clinical classification of coronavirus disease (COVID-19) | 32215691 | S48 | China | English | Cohort Study | Adult | 78 | 44.6 (17.9) | 38 (49) | 40 (51) |
| **Li et al, 2020 [99]** | Characteristics of peripheral blood leukocyte differential counts in patients with COVID-19 | 32114745 | S49 | China | Chinese | Cohort Study | Adult | 10 | 46.5 [36.5-64.3] | 5 (50) | 5 (50) |
| **Li et al, 2020 [96]** | Comparison of epidemic characteristics between SARS in 2003 and COVID-19 in 2020 in Guangzhou | 32159317 | S50 | China | Chinese | Cohort Study | Adult | 346 | 48 [range 3 months-90 yo] | 167 (48) | 179 (52) |
| **Li et al, 2020 [111]** | Comparison of the clinical characteristics between RNA positive and negative patients clinically diagnosed with 2019 novel coronavirus pneumonia | 32087623 | S51 | China | Chinese | Cohort Study | Adult | 31 | 54 | 15 (48) | 16 (52) |
| **Lian et al, 2020 [77]** | Analysis of Epidemiological and Clinical features in older patients with Corona Virus Disease 2019 (COVID-19) out of Wuhan | 32211844 | S52_young | China | English | Cohort Study | Adult (young and middle- | 652 | 41.15 (1.38) | 349 (54) | 303 (46) |

|  |  |  |  |  |  |  | aged < 60 years) |  |  |  |  |
| --- | --- | --- | --- | --- | --- | --- | --- | --- | --- | --- | --- |
| **Lian et al, 2020 [77]** | Analysis of Epidemiological and Clinical features in older patients with Corona Virus Disease 2019 (COVID-19) out of Wuhan | 32211844 | S52_old | China | English | Cohort Study | Adult (elderly >= 60 years) | 136 | 68.28 (7.31) | 58 (43) | 78 (57) |
| **Lin et al, 2020 [40]** | Novel coronavirus pneumonia outbreak in 2019: Computed tomographic findings in two cases | 32056397 | S53 | China | English | Case Series | Adult | 2 | 37 [36-38] | 2 (100) | 0 (0) |
| **Liu et al, 2020 [147]** | Clinical feature of COVID-19 in elderly patients: a comparison with young and middle-aged patients | 32171866 | S54_old | China | English | Cohort Study | Adult (elderly >= 60 years) | 18 | 68.00 [65.25-69.75] | 12 (67) | 6 (33) |
| **Liu et al, 2020 [147]** | Clinical feature of COVID-19 in elderly patients: a comparison with young and middle-aged patients | 32171866 | S54_young | China | English | Cohort Study | Adult (young and middle-aged < 60 years) | 38 | 47 [35.75-51.25] | 19 (50) | 19 (50) |
| **Liu et al, 2020 [153]** | Gross examination of report of a COVID-19 death autopsy | 32198987 | S55 | China | Chinese | Case Study | Adult | 1 | 85 | 1 (100) | 0 (0) |
| **Liu et al, 2020 [90]** | Clinical characteristics of 30 medical workers infected with new coronavirus pneumonia | 32062957 | S56 | China | Chinese | Cohort Study | Adult | 30 | 35 [21-59] | 10 (33) | 20 (67) |
| **Liu et al, 2020 [81]** | Analysis of factors associated with disease outcomes in hospitalized patients with 2019 novel coronavirus disease | 32118640 | S57 | China | English | Cohort Study | Adult | 78 | 38 [33-57] | 39 (50) | 39 (50) |
| **Liu et al, 2020 [74]** | Clinical and biochemical indexes from 2019-nCoV infected patients linked to viral loads and lung injury | 32048163 | S58_adult | China | English | Case Series | Adult | 12 | 63 [53.5-65] | 8 (67) | 4 (33) |
| **Liu et al, 2020 [133]** | Clinical characteristics of novel coronavirus cases in tertiary hospitals in Hubei Province. | 32044814 | S59 | China | English | Cohort Study | Adult | 137 | 57 [range 20-83] | 61 (45) | 76 (55) |
| **Liu et al, 2020 [146]** | Clinical and CT imaging features of the COVID-19 pneumonia: Focus on pregnant women and children | 32171865 | S60_adult | China | English | Cohort Study | Adult | 14 |  | 5 (36) | 9 (64) |
| **Mo et al, 2020 [98]** | Clinical characteristics of refractory COVID-19 pneumonia in Wuhan, China | 32173725 | S61 | China | English | Cohort Study | Adult | 155 | 54 [42-66] | 86 (55) | 69 (45) |
| **Pan et al, 2020 [132]** | Initial CT findings and temporal changes in patients with the novel coronavirus pneumonia (2019-nCoV): a study of 63 patients in Wuhan, China | 32055945 | S62 | China | English | Cohort Study | Adult | 63 | 44.9 (15.2) | 33 (52) | 30 (48) |
| **Peng et al, 2020 [145]** | Clinical characteristics and outcomes of 112 cardiovascular disease patients infected by 2019-nCoV | 32120458 | S63_severe | China | Chinese | Cohort Study | Adult (severe) | 16 | 57.5 [54-63] | 9 (56) | 7 (44) |
| **Peng et al, 2020 [36]** | Clinical characteristics and outcomes of 112 cardiovascular disease patients infected by 2019-nCoV | 32120458 | S63_non severe | China | Chinese | Cohort Study | Adult (non-severe) | 96 | 62 [55-67.5] | 44 (46) | 52 (54) |
| **Qian et al, 2020 [80]** | A COVID-19 Transmission within a family cluster by presymptomatic infectors in China | 32201889 | S64_adult | China | English | Case series | Adult | 7 | 57.5 [44.5-59] | 3 (43) | 4 (57) |
| **Qian et al, 2020 [156]** | Epidemiologic and Clinical Characteristics of 91 Hospitalized Patients with COVID-19 in Zhejiang, China: A retrospective, multi-centre case series | 32181807 | S65 | China | English | Cohort Study | Adult | 91 | 50 [36.5-57] | 37 (41) | 54 (59) |
| **Qu et al, 2020 [61]** | Platelet-to-lymphocyte ratio is associated with prognosis in patients with coronavirus disease-19 | 32181903 | S66 | China | English | Cohort Study | Adult | 30 | 50.5 [36-65] | 16 (53) | 14 (47) |
| **Ren et al, 2020 [165]** | Identification of a novel coronavirus causing severe pneumonia in human - a descriptive study | 32004165 | S67 | China | English | Case Series | Adult | 5 | 52 [49-61] | 3 (60) | 2 (40) |
| **Ruan et al, 2020 [130]** | A case of 2019 novel coronavirus infected pneumonia with twice negative 2019-nCoV nucleic acid testing within 8 days | 32149771 | S68 | China | English | Case study | Adult | 1 | 47 | 0 (0) | 1 (100) |
| **Shi et al, 2020 [44]** | Association of Cardiac Injury With Mortality in Hospitalized Patients With COVID-19 in Wuhan, China | 32211816 | S69 | China | English | Cohort Study | Adult | 416 | 64 [range 21-90] | 205 (49) | 211 (51) |
| **Shi et al, 2020 [51]** | Radiological findings from 81 patients with COVID-19 pneumonia in Wuhan, China: a descriptive study | 32105637 | S70 | China | English | Cohort Study | Adult | 81 | 49.5 (11) | 42 (52) | 39 (48) |
| **Shi et al, 2020 [103]** | Evolution of CT Manifestations in a Patient Recovered from 2019 Novel Coronavirus (2019-nCoV) Pneumonia in Wuhan, China | 32032497 | S71 | China | English | Case Study | Adult | 1 | 42 | 1 (100) | 0 (0) |
| **Silverstein et al, 2020 [91]** | First imported case of 2019 novel coronavirus in Canada, presenting as mild pneumonia | 32061312 | S72 | Canada | English | Case Study | Adult | 1 | 56 | 1 (100) | 0 (0) |
| **Song et al, 2020 [60]** | SARS-CoV-2 induced diarrhoea as onset symptom in patient with COVID-19 | 32139552 | S73 | China | English | Case Study | Adult | 1 | 22 | 1 (100) | 0 (0) |
| **Song et al, 2020 [164]** | Emerging 2019 Novel Coronavirus (2019-nCoV) Pneumonia | 32027573 | S74 | China | English | Cohort Study | Adult | 51 | 49 (16) | 25 (49) | 26 (51) |
| **Spiteri et al, 2020 [31]** | First cases of coronavirus disease 2019 (COVID-19) in the WHO European Region, 24 January to 21 February 2020 | 32156327 | S75 | Europe | English | Cohort Study | Adult | 38 | 42 [range 2-81] | 25 (66) | 13 (34) |
| **Stoecklin et al, 2020 [160]** | First cases of coronavirus disease 2019 (COVID-19) in France: surveillance, investigations and control measures, January 2020 | 32070465 | S76 | France | English | Case Series | Adult | 3 | 31 [30.5-39.5] | 2 (67) | 1 (33) |
| **Sun et al, 2020 [134]** | Epidemiological and Clinical Predictors of COVID-19 | 32211755 | S77 | Singapore | English | Cohort Study | Adult | 54 | 42 [34-54] | 29 (54) | 25 (46) |
| **Sun et al, 2020 [89]** | Evolution of Computed Tomography Manifestations in Five Patients Who Recovered from Coronavirus Disease | 32174054 | S78 | China | English | Case Series | Adult | 5 | 45 [range 20-55] | 2 (40) | 3 (60) |
| 2019 (COVID-19) Pneumonia. |  |  |  |  |  |  |  |  |  |  |  |
| **Tang et al, 2020 [108]** | Abnormal coagulation parameters are associated with poor prognosis in patients with novel coronavirus pneumonia | 32073213 | S79 | China | English | Cohort Study | Adult | 183 | 54.1 (16.2) | 98 (54) | 85 (46) |
| **Tian et al, 2020 [148]** | Characteristics of COVID-19 infection in Beijing | 32112886 | S80 | China | English | Cohort Study | Adult | 262 | 47.5 [range 1-94] | 127 (48) | 135 (52) |
| **Tian et al, 2020 [84]** | Pulmonary Pathology of Early-Phase 2019 Novel Coronavirus (COVID-19) Pneumonia in Two Patients With Lung Cancer | 32114094 | S81_cas e1 | China | English | Case Study | Adult | 1 | 73 | 1 (100) | 0 (0) |
| **Tian et al, 2020 [84]** | Pulmonary Pathology of Early-Phase 2019 Novel Coronavirus (COVID-19) Pneumonia in Two Patients With Lung Cancer | 32114094 | S81_cas e2 | China | English | Case Study | Adult | 1 | 84 | 0 (0) | 1 (100) |
| **Tong et al, 2020 [38]** | Potential Presymptomatic Transmission of SARS-CoV-2, Zhejiang Province, China, 2020 | 32091386 | S82 | China | English | Case Series | Adult | 6 | 23.00 [15.00-41.75] | 3 (50) | 3 (50) |
| **Van Cuong et al, 2020 [154]** | The first Vietnamese case of COVID-19 acquired from China | 32085849 | S83 | Vietnam | English | Case Study | Adult | 1 | 25 | 0 (0) | 1 (100) |
| **Wan et al, 2020 [70]** | Clinical Features and Treatment of COVID-19 Patients in Northeast Chongqing | 32198776 | S84 | China | English | Cohort Study | Adult | 135 | 47 [36-55] | 72 (53) | 63 (47) |
| **Wang et al, 2020 [56]** | Clinical characteristics and therapeutic procedure for four cases with 2019 novel coronavirus pneumonia receiving combined Chinese and Western medicine treatment | 32037389 | S85 | China | English | Case series | Adult | 4 | 47.5 [28.75-63] | 3 (75) | 1 (25) |
| **Wang et al, 2020 [139]** | Clinical Characteristics of 138 Hospitalized Patients with 2019 Novel Coronavirus-Infected Pneumonia in Wuhan, China | 32031570 | S86 | China | English | Cohort Study | Adult | 138 | 56 [42 - 68] | 75 (54) | 63 (46) |
| **Wang et al, 2020 [100]** | Clinical Features of 69 Cases with Coronavirus Disease 2019 in Wuhan, China | 32176772 | S87 | China | English | Cohort Study | Adult | 69 | 42 [35-62] | 32 (46) | 37 (54) |
| **Wang et al, 2020 [45]** | Clinical Outcomes in 55 Patients With Severe Acute Respiratory Syndrome Coronavirus 2 Who Were Asymptomatic at Hospital Admission in Shenzhen, China | 32179910 | S88 | China | English | Cohort Study | Adult | 55 | 49 [range 2-69] | 22 (40) | 33 (60) |
| **Wang et al, 2020 [26]** | The clinical dynamics of 18 cases of COVID-19 outside of Wuhan, China | 32139464 | S89 | China | English | Cohort Study | Adult | 18 | 39 [29-55] | 10 (56) | 8 (44) |
| **Wu et al, 2020 [125]** | Clinical Characteristics of Imported Cases of COVID-19 in Jiangsu Province: A Multicenter Descriptive Study | 32109279 | S90 | China | English | Cohort Study | Adult | 80 | 46.1(15.42) | 39 (49) | 41 (51) |
| **Wu et al, 2020 [85]** | Biological characters analysis of COVID-19 patient accompanied with aplastic anemia | 32145715 | S91 | China | Chinese | Case Study | Adult | 1 | 48 | 1 (100) | 0 (0) |
| **Xie et al, 2020 [82]** | Comparison of different samples for 2019 novel coronavirus detection by nucleic acid amplification tests | 32114193 | S92 | China | English | Case Series | Adult | 9 | 34 [26-45] | 4 (44) | 5 (56) |
| **Xiong et al, 2020 [50]** | Clinical and High-Resolution CT Features of the COVID-19 Infection: Comparison of the Initial and Follow-up Changes | 32134800 | S93 | China | English | Cohort Study | Adult | 42 | 49.5 (14.1) | 25 (60) | 17 (40) |
| **Xu et al, 2020 [73]** | Clinical and computed tomographic imaging features of novel coronavirus pneumonia caused by SARS-CoV-2 | 32109443 | S94 | China | English | Cohort Study | Adult | 50 | 43.9 (16.8) | 29 (58) | 21 (42) |
| **Xu et al, 2020 [22]** | Imaging and clinical features of patients with 2019 novel coronavirus SARS-CoV-2 | 32107577 | S95 | China | English | Cohort Study | Adult | 90 | 50 [range 18-86] | 39 (43) | 51 (57) |
| **Xu et al, 2020 [64]** | Pathological findings of COVID-19 associated with acute respiratory distress syndrome | 32085846 | S96 | China | English | Case Study | Adult | 1 | 50 | 1 (100) | 0 (0) |
| **Xu et al, 2020 [8]** | Clinical findings in a group of patients infected with the 2019 novel coronavirus (SARS-Cov-2) outside of Wuhan, China: retrospective case series | 32075786 | S97 | China | English | Cohort Study | Adult | 62 | 41 [32-52] | 35 (56) | 27 (44) |
| **Xu et al, 2020 [41]** | Clinical features and dynamics of viral load in imported and non-imported patients with COVID-19 | 32179140 | S98_imported | China | English | Cohort Study | Adult | 15 | 35 | 10 (67) | 5 (33) |
| **Xu et al, 2020 [41]** | Clinical features and dynamics of viral load in imported and non-imported patients with COVID-19 | 32179140 | S98_secondary | China | English | Cohort Study | Adult | 17 | 37 | 7 (41) | 10 (59) |
| **Xu et al, 2020 [41]** | Clinical features and dynamics of viral load in imported and non-imported patients with COVID-19 | 32179140 | S98_tertiary | China | English | Cohort Study | Adult | 19 | 53 | 8 (42) | 11 (58) |
| **Yang et al, 2020 [126]** | Clinical characteristics and imaging manifestations of the 2019 novel coronavirus disease (COVID-19):A multi-center study in Wenzhou city, Zhejiang, China | 32112884 | S99 | China | English | Cohort Study | Adult | 149 | 45.11 (13.35) | 81 (54) | 68 (46) |
| **Yang et al, 2020 [9]** | Clinical course and outcomes of critically ill patients with SARS-CoV-2 pneumonia in Wuhan, China: a single-centered, retrospective, observational study | 32105632 | S100 | China | English | Cohort Study | Adult | 52 | 59.7 (13.3) | 35 (67) | 17 (33) |
| **Yao et al, 2020 [42]** | Clinical characteristics and influencing factors of patients with novel coronavirus pneumonia combined with liver injury in Shaanxi region | 32153170 | S101 | China | Chinese | Cohort Study | Adult | 40 | 53.87 (15.84) | 25 (63) | 15 (37) |
| **Yao et al, 2020 [143]** | Epidemiological characteristics of 2019-nCoV infections in Shaanxi, China by February 8, 2020 | 32139462 | S102 | China | English | Cohort Study | Adult | 195 | 44.13 (15.8) | 129 (66) | 66 (34) |
| **Ye et al, 2020 [23]** | Clinical characteristics of severe acute respiratory syndrome coronavirus 2 | 32171867 | S103 | China | English | Case series | Adult | 5 | 31 [30-32] | 2 (40) | 3 (60) |
| **Yoon et al, 2020 [141]** | Chest Radiographic and CT Findings of the 2019 Novel Coronavirus Disease (COVID-19): Analysis of Nine Patients Treated in Korea | 32100485 | S104 | South Korea | English | Cohort Study | Adult | 9 | 54 | 4 (44) | 5 (56) |
| **Young et al, 2020 [79]** | Epidemiologic Features and Clinical Course of Patients Infected With SARS-CoV-2 in Singapore | 32125362 | S105 | Singapore | English | Cohort Study | Adult | 18 | 47 [31-71] | 9 (50) | 9 (50) |
| **Yu et al, 2020 [150]** | A Familial Cluster of Infection Associated With the 2019 Novel Coronavirus Indicating Possible Person-to-Person Transmission During the Incubation Period | 32067043 | S106 | China | English | Case series | Adult | 4 | 72 [68-78.25] | 2 (50) | 2 (50) |
| **Yuan et al, 2020 [55]** | Association of radiologic findings with mortality of patients infected with 2019 novel coronavirus in Wuhan, China | 32191754 | S107 | China | English | Cohort Study | Adult | 27 | 60 [47-69] | 12 (44) | 15 (56) |
| **Zhang et al, 2020 [71]** | CT image of novel coronavirus pneumonia: a case report | 32189175 | S108 | China | English | Case Study | Adult | 1 | 64 | 1 (100) | 0 (0) |
| **Zhang et al, 2020 [27]** | Clinical features of 2019 novel coronavirus pneumonia in the early stage from a fever clinic in Beijing | 32164091 | S109 | China | Chinese | Cohort Study | Adult | 9 | 36 [15-49] | 5 (56) | 4 (44) |
| **Zhang et al, 2020 [34]** | Clinical characteristics of 140 patients infected with SARS-CoV-2 in Wuhan, China | 32077115 | S110 | China | English | Cohort Study | Adult | 140 | 57 [range 25-87] | 71 (51) | 69 (49) |
| **Zhang et al, 2020 [155]** | Epidemiological, clinical characteristics of cases of SARS-CoV-2 infection with abnormal imaging findings. | 32205284 | S111 | China | English | Cohort Study | Adult | 573 | 46.65 (13.83) | 295 (51) | 278 (49) |
| **Zhang et al, 2020 [117]** | High-resolution CT features of 17 cases of Corona Virus Disease 2019 in Sichuan province, China | 32139463 | S112 | China | English | Cohort Study | Adult | 17 | 48.6 [range 23-74] | 8 (47) | 9 (53) |
| **Zhao et al, 2020 [162]** | The characteristics and clinical value of chest CT images of novel coronavirus pneumonia | 32199619 | S113 | China | English | Cohort Study | Adult | 80 | 44 (1.77) | 43 (54) | 37 (46) |
| **Zhao et al, 2020 [78]** | A comparative study on the clinical features of COVID-19 pneumonia to other pneumonias | 32161968 | S114 | China | English | Cohort Study | Adult | 19 | 48 [27-56] | 11 (58) | 8 (42) |
| **Zhou et al, 2020 [48]** | Clinical course and risk factors for mortality of adult inpatients with COVID-19 in Wuhan, China: a retrospective cohort study | 32171076 | S115 | China | English | Cohort Study | Adult | 191 | 56 [46.0-67.0] | 119 (62) | 72 (38) |
| **Zhu et al, 2020 [110]** | Comparison of heart failure and 2019 novel coronavirus pneumonia in chest CT features and clinical characteristics | 32129583 | S116 | China | Chinese | Cohort Study | Adult | 12 | 52 [32-73] | 8 (67) | 4 (33) |
| **Zhu et al, 2020 [58]** | Clinical and CT imaging features of 2019 novel coronavirus disease (COVID-19) | 32142928 | S117 | China | English | Case Series | Adult | 6 | 43 [32-56] | 0 (0) | 6 (100) |

\* mean(sd) or median[Q1-Q3], \*\* COVID-19 National Emergency Response Center, Epidemiology and Case Management Team, Korea Centers for Disease Control and Prevention, Cheongju, Korea et al. 2020

### 2.5. Statistical analysis

For the studies reporting mean and standard deviation (SD) for extracted variables, we computed the median and interquartile ranges (IQR) assuming a normal distribution (i.e., using the formula: IQR ∼ SD*1.35). To test if there is a bias by including the studies for which we computed the median and IQR (i.e., quartiles, Q1 and Q3), we performed a sensitivity analyses in which we calculated the median and IQR under the assumption of right-skewed and left-skewed distribution (see Appendix 2). We compared the results of the different distributions to test the robustness of our findings. Descriptive statistics (median, IQR, n, and %) were used to characterize the studies and patients included as well as the laboratory parameters. Weighted by study sample size, the pooled median and 95% confidence interval (CI) were computed for continuous variables. Normality approximation of the binomial was used to construct an approximate confidence interval (R package metamedian^17^). Welch’s two-sample t-test was employed to test if there are significant differences in the proportion of male and female patients across studies.

Our meta-analysis was structured in two parts. In the first part, we performed meta-analyses of all 148 studies to define the prevalence of comorbidities, clinical signs and symptoms, imaging features, treatments, outcomes, and complications associated with COVID-19. Using the metaprop function of the R package metafor^18^, we calculated the overall prevalence from studies reporting a single prevalence. Our meta-analysis was stratified by patient group (pediatric/neonatal [≤17 years of age], pregnant, and adult COVID-19 patients). Heterogeneity between studies was assessed visually by Forest plots, and analytically by I², tau T^2^, and Cochrane Q. Briefly put, I^2^ describes the percentage of variation across studies that is due to heterogeneity rather than chance^19^: 0% indicates no heterogeneity, whereas 25%, 50%, and 75% indicate low, moderate, and high heterogeneity, respectively. CIs for I² were calculated using the iterative non-central chi-squared distribution method of Hedges and Piggott^20^. Tau (T^2^) represents the absolute value of the true variance (heterogeneity) and is the estimated SD of underlying true effects across studies. Cochran’s Q is the weighted sum of squared differences between individual study effects and the pooled effect across studies, with the weights being those used in the pooling method (i.e., sample size)^21^. The second part comprised meta-analyses to calculate the relative risk (RR) of certain comorbidities, clinical signs and symptoms, imaging features, laboratory parameters, complications, and outcomes in patients with severe vs. those with non-severe disease condition (12 studies) as well as deceased vs. survivors (7 studies). The categorization into severe and non-severe COVID-19 disease was consistent with the groups reported by the reviewed studies (Supplementary Table 2). Owing to our judgment that considerable clinical and statistical heterogeneity exists among the studies (statistical heterogeneity was confirmed by the computed I^2^, T^2^, and Cochrane Q), we calculated pooled RRs with 95% CIs using a random-effects model with inverse-variance weighting (metabin function from R package meta). For continuous outcome data (e.g., age, laboratory parameters, and time from symptoms onset to hospital admission), we estimated the standardized mean difference (SMD) by means of a random-effect models with inverse variance weighting for pooling (metacont function from R package meta). To calculate the SMD, we converted medians, Q1s, and Q3s into means and standard deviations. The SMD, 95% CIs, and p values were reported. We produced Forest plots to visualize the results from the random-effect models (R function: forest). Publication bias was assessed visually by funnel plots (R function: funnel) and analytically by the Egger test (R function: regtest). An Egger test p<0·05 indicates a significant publication bias. All statistical analyses were performed in R (version 3.6.3) for MacOS X (Mojave, 10.14.4) with the packages meta (version 4.11–0) and dmetar (version 0.0.90)^18^. The code used for the analysis and to create figures and tables is provided in our GitHub repository (https://github.com/jutzca/Corona-Virus-Meta-Analysis-2020).

### 2.6. Role of funding source

The funder of the study had no role in study design, data collection, data analysis, data interpretation, or writing of the report. The corresponding author had full access to all the data in the study and had final responsibility for the decision to submit for publication.

## 3. Results

### Study selection and study characteristics

Our systematic literature search yielded 5’049 articles (including articles identified by manual searching). Upon removal of duplicates and exclusion of studies on the basis of their abstracts or following screening their full text, 148 met the inclusion criteria and were considered for the review and meta-analysis (Figure 1)^4,8,28,118–127,29,128–137,30,138–147,31,148–157,32,158–165,33–37,9,38–47,11,48–57,22,58–67,23,68–77,24,78–87,25,88–97,26,98–107,27,108–117^. The inter-rater agreement for study selection was very high (κ = 0·94 [95% CI: 0.91 –0.96], 97.0% agreement [11/ 362 studies with disagreement]). Detailed information on the included studies are provided in Tables 1–3. Included studies were conducted in 15 countries between December 1^st^, 2019 and March 28^th^, 2020 (Supplementary Table 3) and enrolled between 1 and 1’099 patients(median 12.5 [1.00 – 56.75]). The majority of the articles were written in English (123 studies, 83.1%) and the remainder in Chinese (25 studies, 16.9%). We classified studies according to their design^16^: cohort study (76 studies, 51.4%), case study/ report (41 studies, 27.7%), and case series (31 studies, 20.9%). While all studies reported information on demographics (148, 100%), the number of studies reporting information on comorbidities (84 studies, 56.8%), clinical sign and symptoms (130 studies, 87.8%), laboratory parameters (113 studies, 76.4%), imaging features (118 studies, 79.7%), treatments (91,61.5%), outcomes (118 studies, 79.3%), and complications (59 studies, 39.9%) varied markedly.

**Table 2.** Included studies of pregnant women with COVID-19.

| Authors | Title | PMID | Unique study ID | Country | Language | Study type | Study population | Sample size | Age * | Male (%) | Female (%) |
| --- | --- | --- | --- | --- | --- | --- | --- | --- | --- | --- | --- |
| **Chen et al, 2020 [29]** | Pregnant women with new coronavirus infection: a clinical characteristics and placental pathological analysis of three cases | 32114744 | S118 | China | Chinese | Case Series | Pregnant | 3 | 29.6 | 0 (0) | 3 (100) |
| **Chen et al, 2020 [75]** | Chest computed tomography images of early coronavirus disease (COVID-19) | 32162211 | S119 | China | English | Case Study | Pregnant | 1 | 27 | 0 (0) | 1 (100) |
| **Chen et al, 2020 [11]** | Clinical characteristics and intrauterine vertical transmission potential of COVID-19 infection in nine pregnant women: a retrospective review of medical records | 32151335 | S120 | China | English | Case series | Pregnant | 9 | 28 [26-33] | 0 (0) | 9 (100) |
| **Dong et al, 2020 [46]** | Possible Vertical Transmission of SARS-CoV-2 From an Infected Mother to Her Newborn | 32215581 | S121_pregnant | China | English | Case Study | Pregnant | 1 | 29 | 0 (0) | 1 (100) |
| **Liao et al, 2020 [122]** | Chest CT Findings in a Pregnant Patient with 2019 Novel Coronavirus Disease | 32212578 | S122 | China | English | Case Study | Pregnant | 1 | 25 | 0 (0) | 1 (100) |
| **Liu et al, 2020 [146]** | Clinical and CT imaging features of the COVID-19 pneumonia: Focus on pregnant women and children | 32171865 | S60_pregnant | China | English | Cohort Study | Pregnant | 16 | 30 [26-35] | 0 (0) | 16 (100) |
| **Wang et al, 2020 [123]** | A case of 2019 Novel Coronavirus in a pregnant woman with preterm delivery | 32119083 | S123 | China | English | Case study | Pregnant | 1 | 28 | 0 (0) | 1 (100) |
| **Wang et al, 2020 [152]** | A case report of neonatal COVID-19 infection in China | 32161941 | S124_pregnant | China | English | Case study | Pregnant | 1 | 34 | 0 (0) | 1 (100) |
| **Wen et al, 2020 [124]** | A patient with SARS-CoV-2 infection during pregnancy in Qingdao, China | 32198004 | S125 | China | English | Case study | Pregnant | 1 | 31 | 0 (0) | 1 (100) |
| **Xia et al, 2020 [136]** | Emergency Caesarean delivery in a patient with confirmed coronavirus disease 2019 under spinal anaesthesia | 32192711 | S126 | China | English | Case Study | Adult | 1 | 27 | 0 (0) | 1 (100) |

\* mean(sd) or median[Q1-Q3]

**Table 3.** Included studies of pregnant women with COVID-19.

| Authors | Title | PMID | Unique study ID | Country | Language | Study type | Study population | Sample size | Age * | Male (%) | Female (%) |
| --- | --- | --- | --- | --- | --- | --- | --- | --- | --- | --- | --- |
| **Cai et al, 2020 [65]** | First case of 2019 novel coronavirus infection in children in Shanghai | 32102141 | S127 | China | Chinese | Case Study | Pediatric | 1 | 7 | 1 (100) | 0 (0) |
| **Chan et al, 2020 [4]** | A familial cluster of pneumonia associated with the 2019 novel coronavirus indicating person-to-person transmission: a study of a family cluster | 31986261 | S6_pediatric | China | English | Case series | Pediatric | 1 | 10 | 1 (100) | 0 (0) |
| **Chen et al, 2020 [101]** | First case of severe childhood novel coronavirus pneumonia in China | 32135586 | S128 | China | Chinese | Case Study | Pediatric | 1 | 1.1 | 1 (100) | 0 (0) |
| **Cui et al, 2020 [33]** | A 55-Day-Old Female Infant Infected With 2019 Novel Coronavirus Disease: Presenting With Pneumonia, Liver Injury, and Heart Damage | 32179908 | S129 | China | English | Case study | Neonatal | 1 | 55 (days) | 0 (0) | 1 (100) |
| **Dong et al, 2020 [46]** | Possible Vertical Transmission of SARS-CoV-2 From an Infected Mother to Her Newborn | 32215581 | S121_pediatric | China | English | Case Study | Neonatal | 1 | 0 | 0 (0) | 1 (100) |
| **Dong et al, 2020 [92]** | Epidemiological Characteristics of 2143 Pediatric Patients With 2019 Coronavirus Disease in China | DOI: 10.1542/peds.2020-0702 | S130 | China | English | Cohort Study | Pediatric | 731 | 10 [2-13] | 420 (57) | 311 (43) |
| **Fan et al, 2020 [158]** | Anal swab findings in an infant with COVID-19 | DOI : 10.1002/ped4.12186 | S131 | China | English | Case Study | Neonatal | 1 | 0.25 | 0 (0) | 1 (100) |
| **Feng et al, 2020 [87]** | Analysis of CT features of 15 children with 2019 novel coronavirus infection | 32061200 | S132 | China | Chinese | Case series | Pediatric | 15 | 7 [range 4-14] | 5 (33) | 10 (67) |
| **Ji et al, 2020 [135]** | Clinical features of pediatric patients with COVID-19: a report of two family cluster cases | 32180140 | S133 | China | English | Case Series | Pediatric | 2 | 12.0 [10.5-13.5] | 2 (100) | 0 (0) |
| **Le et al, 2020 [24]** | The first infant case of COVID-19 acquired from a secondary transmission in Vietnam | 32213326 | S134 | Vietnam | English | Case Study | Neonatal | 1 | 0.25 | 0 (0) | 1 (100) |
| **Liu et al, 2020 [74]** | Clinical and biochemical indexes from 2019-nCoV infected patients linked to viral loads and lung injury | 32048163 | S58_pediatric | China | English | Case Study | Pediatric | 1 | 10 | 1 (100) | 0 (0) |
| **Liu et al, 2020 [86]** | Detection of Covid-19 in Children in Early January 2020 in Wuhan, China | 32163697 | S135 | China | English | Case Series | Pediatric | 6 | 3 [3-3.75] | 2 (33) | 4 (67) |
| **Liu et al, 2020 [146]** | Clinical and CT imaging features of the COVID-19 pneumonia: Focus on pregnant women and children | 32171865 | S60_pediatric | China | English | Cohort Study | Pediatric | 4 | 3.0 [0.7-6.0] | 2 (50) | 2 (50) |
| **Lu et al, 2020 [37]** | SARS-CoV-2 Infection in Children | 32187458 | S136 | China | English | Cohort Study | Pediatric | 171 | 6.7 [2-9.8] | 104 (61) | 67 (39) |
| **Park et al, 2020 [163]** | First Pediatric Case of Coronavirus Disease 2019 in Korea | 32193905 | S137 | South Korea | English | Case Study | Pediatric | 1 | 10 | 0 (0) | 1 (100) |
| **Qian et al, 2020 [80]** | A COVID-19 Transmission within a family cluster by presymptomatic infectors in China | 32201889 | S64_pediatric | China | English | Case study | Pediatric (asymptomatic) | 1 | 1.1 | 0 (0) | 1 (100) |
| **Sun et al, 2020 [116]** | Clinical features of severe pediatric patients with coronavirus disease 2019 in Wuhan: a single center's observational study | 32193831 | S138 | China | English | Case series | Pediatric | 8 | 10.2 [5.04-13.54] | 6 (75) | 2 (25) |
| **Tang et al, 2020 [43]** | Detection of Novel Coronavirus by RT-PCR in Stool Specimen from Asymptomatic Child, China | 32150527 | S139 | China | English | Case Study | Pediatric | 1 | 10 | 1 (100) | 0 (0) |
| **Wang et al, 2020 [83]** | SARS-CoV-2 infection with gastrointestinal symptoms as the first manifestation in a neonate | 32204755 | S140 | China | Chinese | Case Study | Neonatal | 1 | 19 (days) | 1 (100) | 0 (0) |
| **Wang et al, 2020 [152]** | A case report of neonatal COVID-19 infection in China | 32161941 | S124_pediatric | China | English | Case study | Neonatal | 1 | 0 | 1 (100) | 0 (0) |
| **Wang et al, 2020 [107]** | Clinical analysis of 31 cases of 2019 novel coronavirus infection in children from six provinces (autonomous region) of northern China | 32118389 | S141 | China | Chinese | Cohort Study | Pediatric | 31 | 7.1 [0.6-17] | 15 (48) | 16 (52) |
| **Wei et al, 2020 [53]** | Novel Coronavirus Infection in Hospitalized Infants Under 1 Year of Age in China | 32058570 | S142 | China | English | Case Series | Neonatal | 9 | 0.583 [0.33-0.75] | 2 (22) | 7 (78) |
| **Xia et al, 2020 [151]** | Clinical and CT features in pediatric patients with COVID-19 infection: Different points from adults | 32134205 | S143 | China | English | Cohort Study | Pediatric | 20 | 2 [range 1 day-14 years 7 months] | 13 (65) | 7 (35) |
| **Xu et al, 2020 [35]** | Characteristics of pediatric SARS-CoV-2 infection and potential evidence for persistent fecal viral shedding | PMCID: PMC7095102 | S144 | China | English | Case Series | Pediatric | 10 | 6.63 [2.17-13.4] | 6 (60) | 4 (40) |
| **Zeng et al, 2020 [97]** | First case of neonate infected with novel coronavirus pneumonia in China | 32065520 | S145 | China | Chinese | Case Study | Neonatal | 1 | 17 (days) | 1 (100) | 0 (0) |
| **Zhang et al, 2020 [145]** | 2019-novel coronavirus infection in a three-month-old baby | 32043842 | S146 | China | Chinese | Case study | Neonatal | 1 | 0.25 | 0 (0) | 1 (100) |
| **Zheng et al, 2020 [129]** | Clinical Characteristics of Children with Coronavirus Disease 2019 in Hubei, China | 32207032 | S147 | China | English | Cohort Study | Pediatric | 25 | 3 [2-9] | 14 (56) | 11 (44) |
| **Zhou et al, 2020 [121]** | Clinical features and chest CT findings of coronavirus disease 2019 in infants and young children | 32204756 | S148 | China | Chinese | Case Series | Pediatric | 9 | 1 [range 7 months-3 years] | 4 (44) | 5 (56) |

\* mean(sd) or median[Q1-Q3]

In terms of study population, 114 studies included only adult participants, 6 only pregnant women, 22 only children and neonates, and 6 included mixed cohorts. Of the total 12’149 patients included, 6’410 (52.8%) were male and 5’739 female (47.2%, Figure 2A and 2B). The median age of adult (11’058 patients, 91.0%), pregnant (35 patients, 0.3%), and pediatric (1’056 patients, 8.7%; including neonates) patients was 47.0 years [35.0–65.3] (Figure 3A), 30.0 [26.0–33.0] Figure 3B, and 10.0 [2.0–13.0] (Figure 3C), respectively. Approximately 7.8% (297/ 3’822 patients) were reported to be asymptomatic and 7.7% (617/ 8’047) died during hospitalization due to complications related to the infection with SARS-CoV-2. With the exception of one 10-month old child, all deaths were non-pregnant adult COVID-19 patients.

**Figure 2.**
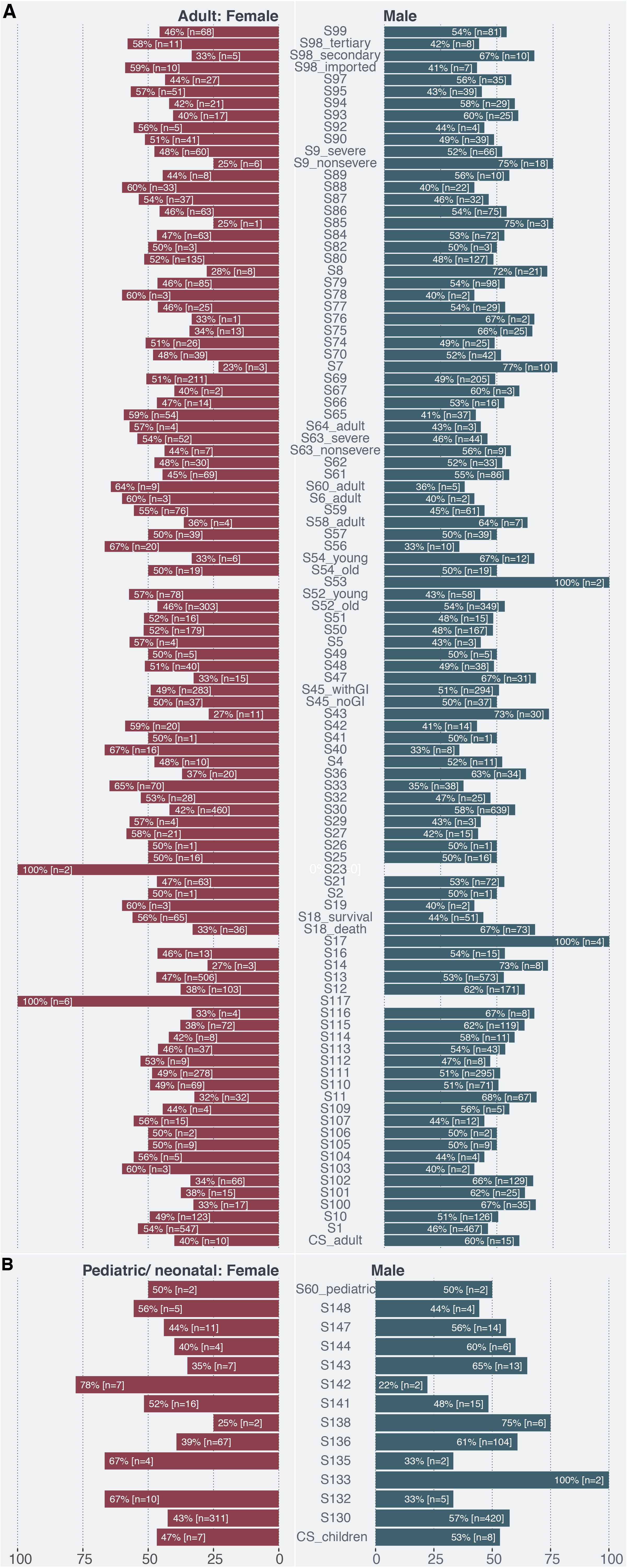
Proportion of female and male patients in adult (A) and pediatric/neonatal cohort (B). All case studies/ reports were pooled together for visualization (CS_adult, and CS_children [pediatric/neonatal]). The key to the study identifier can be found in Tables 1 (adults) and Table 3 (children).

**Figure 3.**
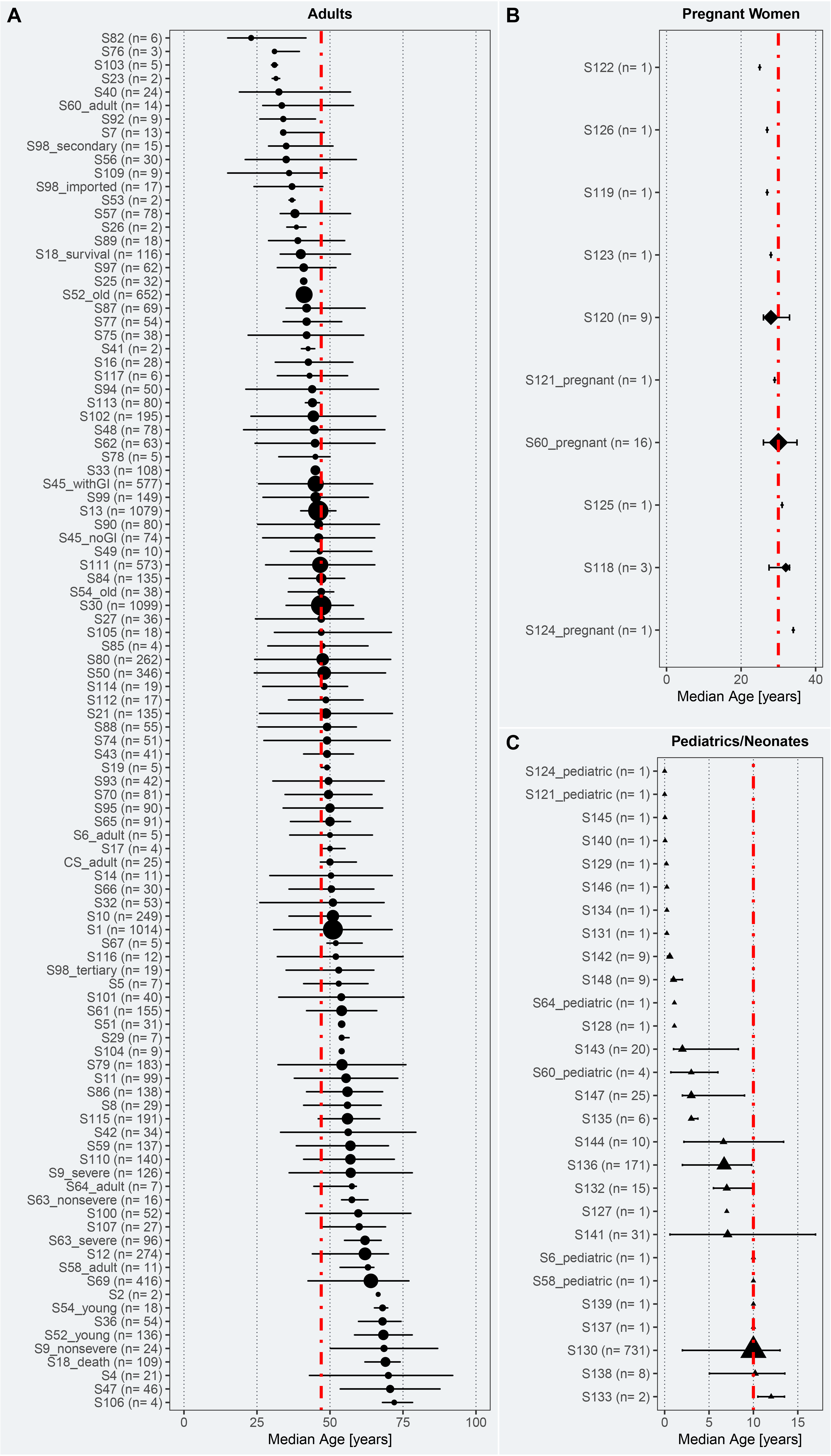
Age of adult (A), pregnant (B), and pediatric/neonatal (C) COVID-19 patients included in eligible studies. Median age and interquartile ranges (IQR) are represented by the midpoints and error bars, respectively. The studies have been sorted by patients’ median age in years. The size of the midpoint (circle, square, triangle) indicates the study sample size. The red line indicates the pooled median age of the respective cohort. All adult case studies/ reports (CS_adult) were pooled for the visualization reasons. The key to the study identifier can be found in Table 1 (adults), Table 2 (pregnant women), and Table 3 (children).

### Adult patients

Higher proportions of male than female patients were reported to be infected with SARS-CoV-2 (t = 2.678, df = 202, p-value = 0.008; Figure 2A) across all studies. Comorbidities were present in ∼31% of the adult patients (2’329/ 7’608), with hypertension being the most prevalent one (1’352/ 6’460 patients, 20.93%), followed by heart failure (37/ 354 patients, 10.5%), diabetes mellitus (678/ 6’535 patients, 10.4%), and coronary heart disease (194/ 2’388 patients, 8.5%) (Figure 4A, Table 4, Supplementary Figure 1). The most frequent clinical signs and symptoms were fever (6’955/ 8’859 patients, 78.5%), cough (4’778/ 8’885 patients, 53.8%), and fatigue (1’996/ 7’980 patients, 25.0%) (Figure 4B, Table 4). A little over five percent of the adult COVID-19 patients were asymptomatic (148/ 2’749 patients, 5.4%). Over 6’969 patients (89.6%) had abnormal CT imaging features. The most common patterns of CT abnormalities were indicating pneumonia (unilateral or bilateral; 6’620/ 7’917 patients, 83.6%), including air bronchogram (264/ 523 patients, 50.5%), and ground-glass opacity (GGO) with consolidation (153/ 323 patients, 47.4%) and without (2’446/ 5’591 patients, 43.8%) (Table 4, Supplementary Figure 2). In terms of laboratory parameters, inflammatory markers, such as interleukin 6 (22 pg/mL [4.68–51.8]), and erythrocyte sedimentation rate (32.5 mm/h [17.3–53.8]) were elevated across the adult population. Moreover, markers of coagulation, namely d-dimer (0.5 µg/mL [0.3–1.08]), fibrinogen (4.5 g/L [3.66–5.1]), and cell damage were also elevated (i.e., lactate dehydrogenase, U/L; 213 [173–268]). An overview of all laboratory parameters is provided in Supplementary Table 4. As shown in Figure 4D, the most common treatments were antivirals (4’475/6’068, patients, 73.8%), oxygen therapy (1’300/ 1’872 patients, 69.4%), and antibiotics (2’518/ 4’825 patients, 52.2%). Detailed information on all treatments is provided in in Table 4. Eight percent (616/7’727 patients) of the adults died during the hospitalization due to complications related to COVID-19. Amongst the survivors (7’111/ 7’727 patients, 92.0%), a total of 3’025 (68.7%) remained hospitalized, 1’751 (32.4%) were discharged, and 1’012 (27.1%) reportedly recovered (Figure 4C, Table 4). Important to note, for some patients it was stated that they both, recovered and were discharged (i.e., one patient can fall in multiple categories). The median duration between symptoms onset and hospitalization was 8 days [7 – 9.5]. A total of 195 (6.8%) patients were admitted to the intensive care unit (ICU). The most frequently reported complications associated with COVID-19 were pneumonia (1’032/ 1’489 patients, 69.2%), respiratory failure (141/ 413 patients, 34.1%), acute cardiac injury (242/ 1’250 patients, 19.4%), and ARDS (759/ 5’122 patients, 14.8%), (Figure 4D, Table 4).

**Table 4:**
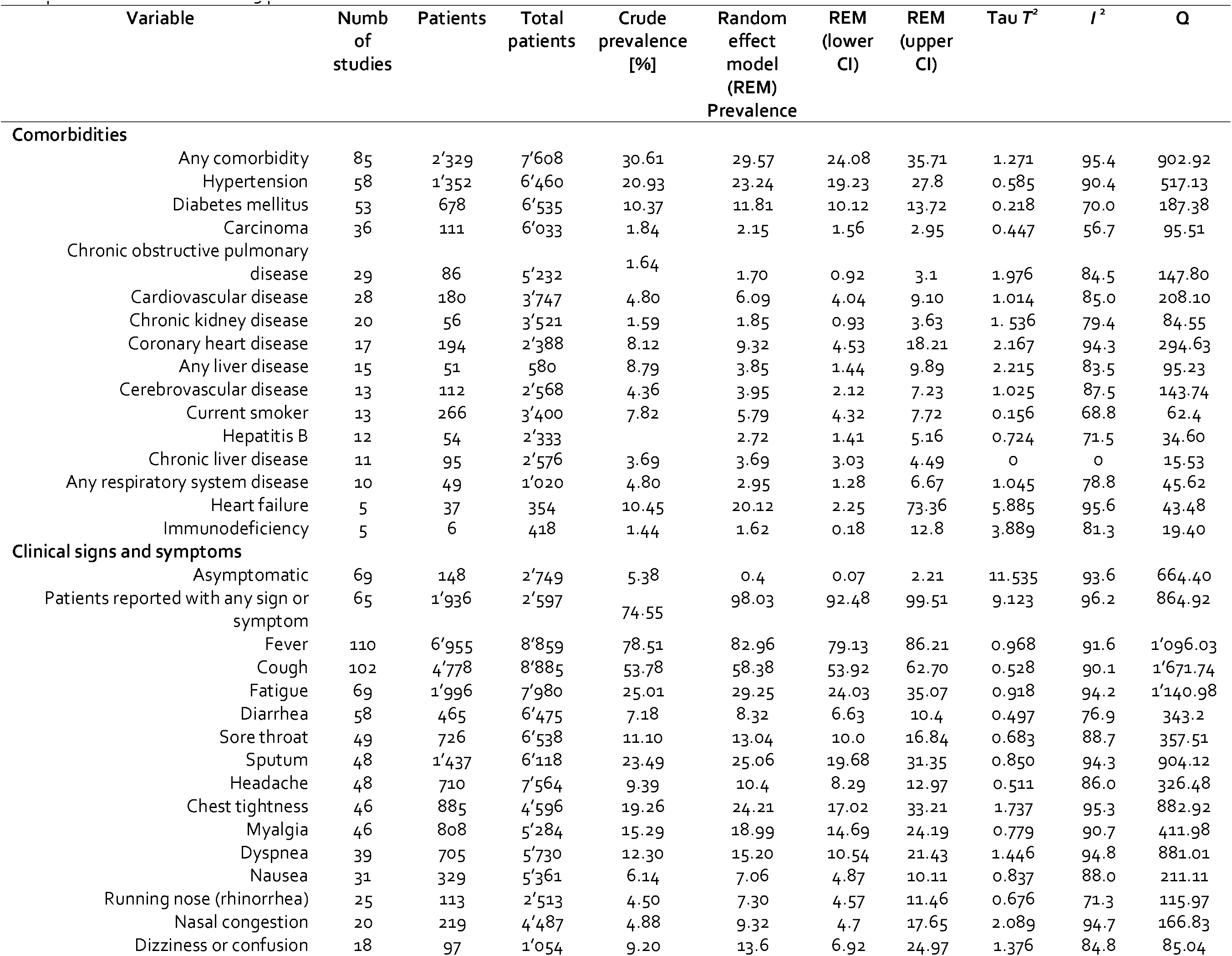
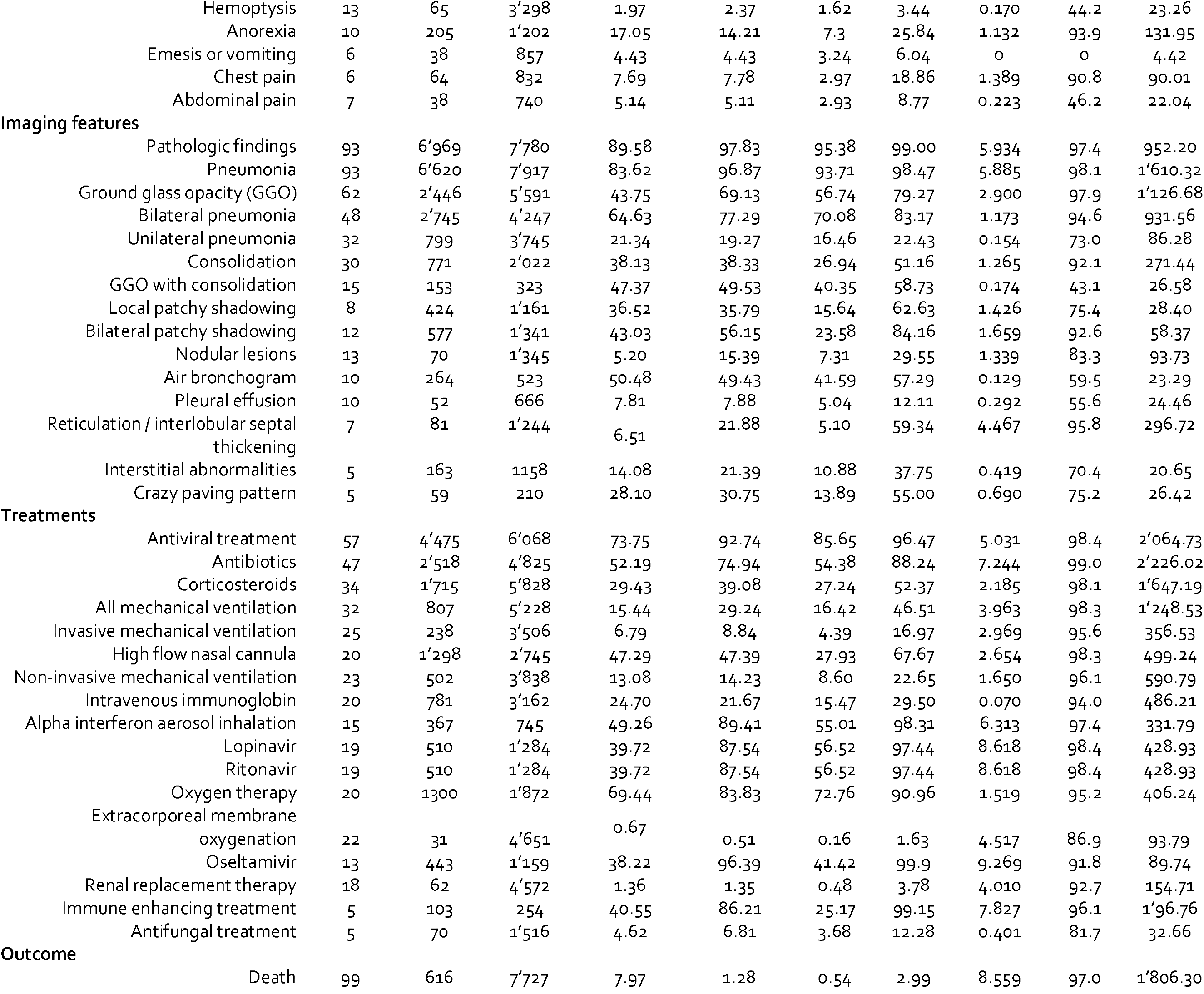
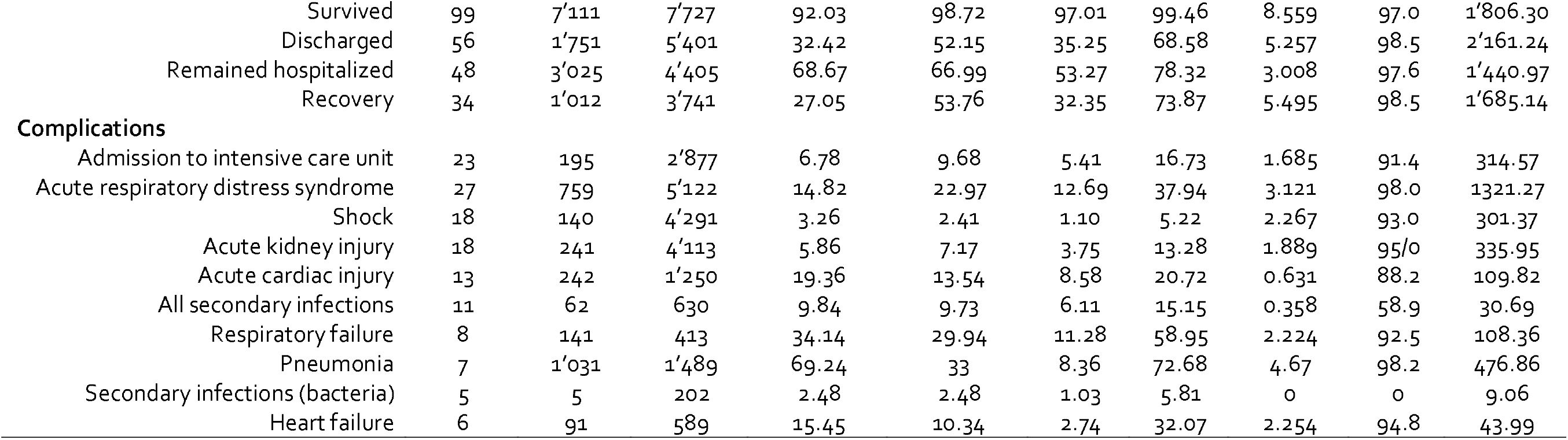
Summary for random effects model for prevalence of comorbidities, clinical signs and symptoms, imaging features, treatments, outcome and complications in adult CoVID-19 patients.

**Figure 4.**
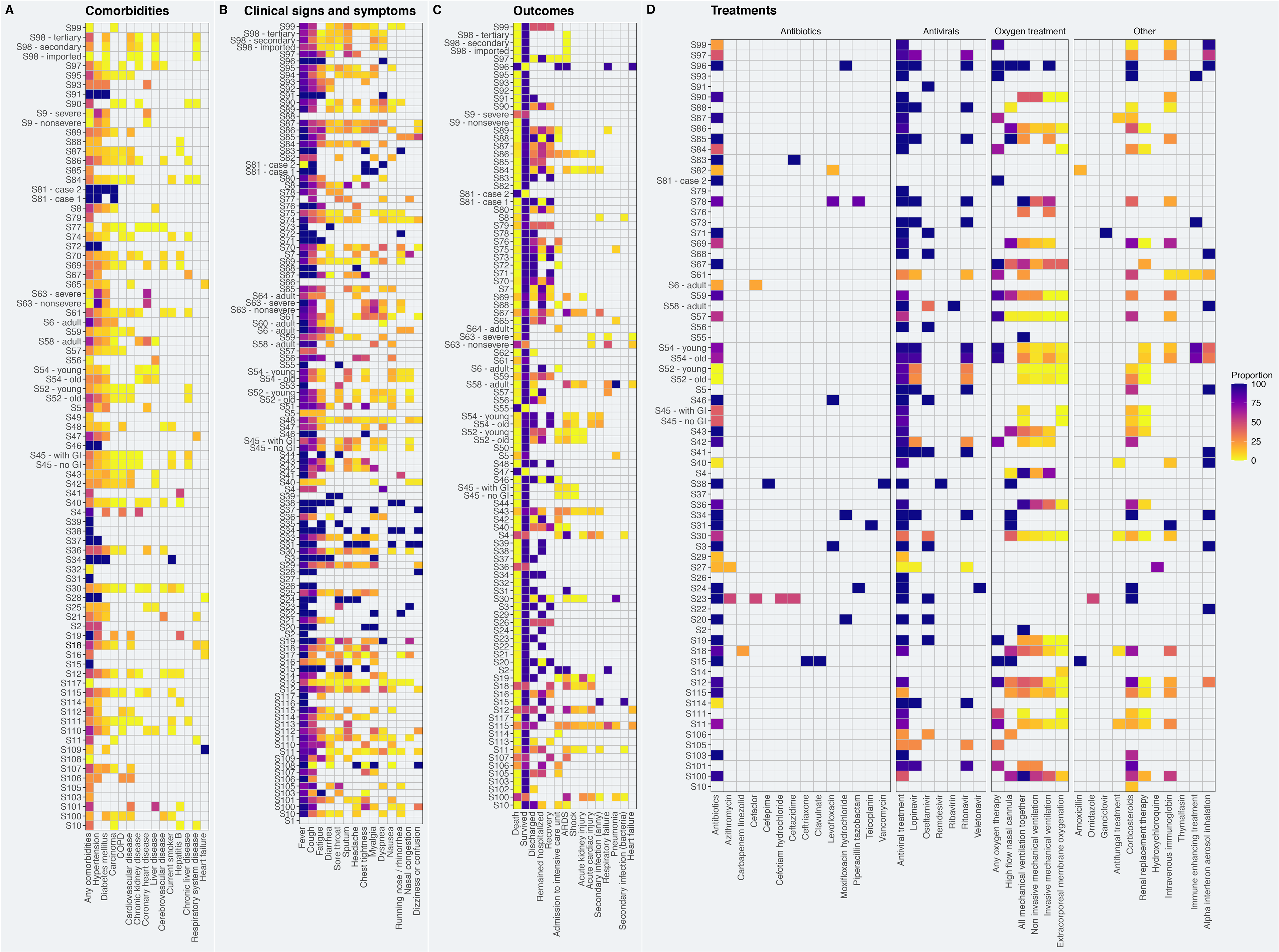
Comorbidities (A), Clinical signs and symptoms (B), outcomes (C), and treatments administered (D) to adult COVID-19 patients. The colors indicated the proportion of patients (%, 0 = yellow, 100 = dark purple). Note: Missing values are colored in white.

### Pregnant woman

Studies investigating the effect of COVID-19 in pregnant women reported that only five pregnant women had any history of comorbidities. Hypothyroidism, allergies, or influenza were reported each for one pregnant woman (Supplementary Table 5). Fever (25/ 35 patients, 71.4%), cough (12/29 patients, 41.4%), and myalgia (3/ 9 patients, 33.3%) were the three most common symptoms observed in pregnant women that were infected with SARS-CoV-2 (Supplementary Figure 3, Supplementary Table 5). Abnormal CT features were evident in 88.6% (31/ 35 patients) of pregnant women diagnosed with COVID-19. Pneumonia (unilateral or bilateral, 31/ 35 patients, 88.6%), GGO (29/ 34 patients, 85.3%), and consolidation (8/ 16 patients, 50.0%) were among the most common patterns of CT abnormalities (Supplementary Figure 4, Supplementary Table 5). Inflammatory markers, such as C-reactive protein (19.25 mg/L [12.35–25.7]), procalcitonin (0.187 ng/mL), and neutrophil count (9.14 × 10^9^/L) were elevated in this patient population. Along this line, lactate dehydrogenase concentrations were increased (544 U/L) reflecting cellular damage. An overview of all laboratory parameters is provided in Supplementary Table 4. Moreover, antibiotics (14/ 14 patients, 100.0%), antivirals (11/ 14 patients, 78.6%) and oxygen therapy (high flow nasal cannula; 3/ 12 patients, 25.0%) were used to treat pregnant COVID-19 patients (Supplementary Table 5). None of the pregnant COVID-19 patients died. Lastly, one patient was admitted to the ICU (Supplementary Table 5).

### Pediatric and Neonatal Patients

Similar to the adult cohort, the proportion between female and male patients were comparable in the pediatric/neonatal cohort (t = 1.169, df = 26, p-value = 0.253; Figure 2B). Fourteen percent of the children and neonates were asymptomatic (149/ 1’054). With the exception of two children, no comorbidities were reported for any of the pediatric or neonatal patients (Supplementary Table 6). Similar to the adult and pregnant COVID-19 patients, children and neonates frequently presented with fever (170/ 320 patients, 53.1%), cough (149/ 311 patients, 47.9%), and sputum (14/ 51 patients, 27.5%) (Supplementary Figure 6 and Supplementary Table 6). Sixty-five percent of the pediatric and neonatal patients presented with CT abnormalities, including pneumonia (194/ 298 patients), GGO (108/ 278 patients, 38.9%), and local patchy shadowing (52/ 223 patients, 23.3%) (Supplementary Figure 7, Supplementary Table 6). An overview of all laboratory parameters is provided in Supplementary Table 7. As the reference values vary considerably within the pediatric/neonatal patient population, the results of the laboratory parameters have to be interpreted with caution. In terms of treatment, children and neonates received antibiotics (31/ 43 patients, 72.1%), oxygen therapy through high flow nasal cannula (5/ 9 patients, 55.6%), and alpha interferon aerosol inhalation therapy (31/52, 59.6%) to treat COVID-19 and its complications (Supplementary Figure 8, Supplementary Table 6). With the exception of a 10-month-old child that died four weeks after admission of multi-organ failure, all children survived. Less than 30% remained hospitalized (90/ 293 patients), 74.5% were discharged (216/ 290 patients) and 87.4% reportedly recovered (236/ 270 patients) (Supplementary Figure 9, Supplementary Table 6). The median duration between symptoms onset and hospitalization was 6 days [4.0–8.5]. Fifteen percent (6/ 39 patients) had to be admitted to the ICU. Complications associated with COVID-19 comprised pneumonia (16/ 26 patients, 61.5%), secondary bacterial infection (12/ 21 patients, 57.1%), and respiratory failure (10/ 33 patients, 30.3%) (Supplementary Table 6).

### Non-severe vs. Severe

Twelve studies (2’596 patients) provided separate data for patients with a severe (500 patients, 19.3%) and non-severe disease status (2’096, 80.7%). No differences regarding sex were found between severe (t = 0.604, df = 16.645, p-value = 0.554; male: 278 patients [55.6%] and female: 210 patients [42.0%]; unknown sex: 12 patients [2.4%]) and non-severe disease status group (t = 0.217, df = 16.393, p-value = 0.831; male: 1’059 patients [50.5%] and female: 925 patients [49.5%]) (Supplementary Figure 10). In terms of age, patients with non-severe COVID-19 were significantly younger (median age in years = 45.0 [34.0–57.0]) than those with a severe disease progression (61.4 [44.5–75.5], Figure 5). Our meta-analysis revealed that older age (SMD: 0.68 [0.40–0.97]; p < 0.001), being male (RR = 1.11 [1.01–1.22]; p = 0.039), and preexisting comorbidities (RR = 2.11 [1.02–4.35], p = 0.046) were associated with a higher risk of increased disease severity. Specifically, hypertension (RR = 2.15 [1.64–2.81], p < 0.001), diabetes mellitus (RR = 2.56 [1.50–4.39], p = 0.005), any heart condition (RR = 4.09 [2.45–6.84], p < 0.001), and chronic obstructive pulmonary disease (COPD, RR = 5.10 [3.08–8.45], p < 0.001) (Figure 6, Table 5) were associated with worse outcome (i.e., severe disease). To test if the increased risk of heart conditions is attributable to the study that has classified their patients into severe and non-severe based on the presence or absence of cardiac injuries, we conducted a sensitivity analysis excluding these studies^44^. The risk of any heart condition remained significantly elevated in the severe disease patient cohort (RR = 3.87 [1.85 – 8.11], p = 0.005). Numerous laboratory parameters were significantly different between the non-severe and severe patient cohorts. Patients with severe disease status presented with decreased levels of albumin (SMD = 1.60 [−2.97 – (−0.24)]; p = 0.022), hemoglobin (SMD = –0.23 [−0.41-(−0.06)]; p = 0.001), and thrombocytes (SMD = –0.57 [−0.68-(−0.45)]; p < 0.001) in comparison to patients with non-severe disease status. Additionally, C-reactive protein (SMD = 1.47 [0.88–2.07]; p < 0.001), lactate dehydrogenase (SMD = 1.71 [1.08–2.34]; p < 0.001), and aspartate transaminase levels (SMD = 0.85 [0.61–1.09]; p < 0.001) were elevated in patients with severe disease status. In terms of complications, patients with severe COVID-19 disease were at an elevated risk of developing ARDS (RR = 10.59 [2.44–46.01], p = 0.014, Figure 6). The heterogeneity between the studies varied substantially (Table 5). Publication bias, measured by means of the Egger’s test, was only evident in three analyses. However, Egger’s test may lack the statistical power to detect bias when the number of studies is small (i.e., fewer than 10) as we only included 4–8 studies.

**Table 5:** Results of meta-analyses for patients with severe and non-severe disease outcome as well as survivors and non-survivors.

|  | Number<br>of<br>Studies | Number of<br>events/ Number<br>of severe | Number of<br>events/ Number<br>of non-severe | RR [95% CI] | p-value | Tau<sup>2</sup> | I<sup>2</sup> | Cochranes<br>Q | Egger's test<br>(p-value) |
| --- | --- | --- | --- | --- | --- | --- | --- | --- | --- |
| **Severe (cases) vs non-severe CoVID-19 disease (controls)** |  |  |  |  |  |  |  |  |  |
| **Demographics** |  |  |  |  |  |  |  |  |  |
| Sex: male | 10 | 278/488 | 1059/1987 | 1.11 [1.01-1.22] | 0.039 | 0.004 | 0% | 7.67 | 0.763 |
| Sex: female | 10 | 210/488 | 925/1987 | 0.95 [0.82-1.10] | 0.450 | 0.006 | 18.6% | 11.05 | 0.395 |
| Age | 11 | 487 | 2059 | SMD: 0.68 [0.40-0.97] | <0.001 | 0.154 | 81.8% | 55.05 | 0.012 |
| **Comorbidities** |  |  |  |  |  |  |  |  |  |
| Any comorbidity | 4 | 167/307 | 291/1205 | 2.11 [1.02-4.35] | 0.046 | 0.160 | 79.8% | 14.86 | 0.122 |
| Hypertension | 8 | 158/429 | 292/1734 | 2.15 [1.64-2.81] | <0.001 | 0.018 | 35.8% | 10.91 | 0.664 |
| Diabetes mellitus | 7 | 84/427 | 127/1720 | 2.56 [1.50-4.39] | 0.005 | 0.038 | 49.7% | 11.92 | 0.279 |
| Any heart condition | 7 | 64/427 | 58/1720 | 4.09 [2.45-6.84] | <0.001 | 0.032 | 22.7% | 7.76 | 0.548 |
| COPD | 6 | 23/403 | 15/1594 | 5.10 [3.08-8.45] | <0.001 | 0 | 0% | 1.59 | 0.034 |
| Carcinoma | 5 | 15/345 | 19/1512 | 3.13 [0.63-15.64] | 0.120 | 0.696 | 42.9% | 7.00 | 0.339 |
| **Symptoms and signs** |  |  |  |  |  |  |  |  |  |
| Fever | 8 | 399/462 | 1588/1847 | 1.02 [0.99-1.06] | 0.187 | <0.0001 | 41.3% | 11.92 | 0.644 |
| Fatigue | 8 | 199/462 | 611/1847 | 1.21 [0.99-1.48] | 0.059 | 0.004 | 46.0% | 12.95 | 0.011 |
| Myalgia | 5 | 53/318 | 237/1454 | 1.01 [0.66-1.56] | 0.929 | <0.0001 | 20.7% | 5.04 | 0.702 |
| Headache | 7 | 47/404 | 187/1765 | 1.14 [0.94-1.39] | 0.146 | <0.0001 | 0.0% | 1.65 | 0.625 |
| Cough | 8 | 290/462 | 1051/1847 | 1.14 [1.02-1.27] | 0.026 | 0.006 | 15.1% | 8.25 | 0.633 |
| Sputum | 6 | 85/385 | 384/1549 | 1.05 [0.79-1.39] | 0.460 | <0.0001 | 14.8% | 5.87 | 0.8731 |
| Dyspnea | 6 | 91/207 | 56/587 | 4.67 [0.99-21.91] | 0.050 | 1.156 | 76.2% | 21.03 | 0.148 |
| Sore throat / Pharyngalgia | 6 | 41/358 | 182/1549 | 1.40 [0.62-3.17] | 0.337 | 0.218 | 50.9% | 10.19 | 0.831 |
| Diarrhea | 6 | 41/403 | 77/1594 | 1.76 [0.72-4.32] | 0.164 | 0.296 | 53.7% | 10.80 | 0.384 |
| **Treatment** |  |  |  |  |  |  |  |  |  |
| Antibiotics | 4 | 254/309 | 743/1410 | 1.63 [0.67-3.96] | 0.177 | 0.285 | 93.5% | 45.93 | 0.807 |
| Antiviral treatment | 6 | 249/347 | 888/1526 | 1.05 [0.90-1.22] | 0.490 | 0.011 | 77.7% | 22.45 | 0.604 |
| Corticosteroids | 5 | 200/345 | 416/1512 | 2.26 [1.32-3.87] | 0.014 | 0.174 | 93.7% | 63.66 | 0.211 |
| **Imaging features (CT)** |  |  |  |  |  |  |  |  |  |
| Pathological findings | 7 | 400/416 | 1372/1631 | 1.06 [0.96-1.18] | 0.192 | 0.009 | 90.1% | 60.32 | 0.085 |
| Pneumonia | 5 | 373/389 | 1290/1539 | 1.05 [0.94-1.18] | 0.299 | 0.008 | 92.1% | 50.58 | 0.176 |
| **Complications** |  |  |  |  |  |  |  |  |  |
| Acute respiratory distress symptom (ARDS) | 4 | 117/331 | 65/1457 | 10.59 [2.44-46.01] | 0.014 | 0.606 | 84.1% | 18.90 | 0.067 |
| Acute kidney injury | 4 | 16/331 | 8/1457 | 6.60 [0.37-116.33] | 0.128 | 2.075 | 65.0% | 8.56 | 0.909 |
| **Laboratory parameter** | **Number of studies** | **Number of severe** | **Number of non-severe** | **SMD [95% CI]** | **p-value** | **Tau<sup>2</sup>** | **I<sup>2</sup>** | **Cochranes Q** | **Egger's test (p-value)** |
| Albumin | 3 | 131 | 511 | -1.60 [-2.97 - (-0.24)] | 0.022 | 1.385 | 96% | 50.01 | 0.790 |
| Alanine aminotransferase (ALT) | 6 | 184 | 695 | 0.27 [0.06-0.47] | 0.011 | 0.014 | 22.1% | 6.42 | 0.545 |
| Aspartate transaminase (AST) | 6 | 184 | 695 | 0.85 [0.61-1.09] | <0.001 | 0.031 | 36.5% | 7.88 | 0.942 |
| Creatinine | 6 | 205 | 794 | 0.59 [0.12-1.07] | 0.015 | 0.298 | 87.3% | 39.30 | 0.501 |
| C-reactive protein (CRP) | 6 | 227 | 774 | 1.47 [0.88-2.07] | <0.001 | 0.487 | 91.2% | 56.50 | 0.296 |
| D-Dimer | 4 | 143 | 361 | 0.55 [0.22-0.89] | 0.001 | 0.066 | 59.4% | 7.39 | 0.632 |
| Hemoglobin | 6 | 342 | 1618 | -0.23 [-0.41 - (-0.06)] | 0.001 | 0.016 | 37.2% | 7.96 | 0.927 |
| Lactate dehydrogenase (LDH) | 4 | 93 | 279 | 1.71 [1.08-2.34] | <0.001 | 0.294 | 77.3% | 13.20 | 0.599 |
| Leucocytes | 7 | 412 | 1676 | 0.49 [-0.24-1.21] | 0.187 | 0.905 | 97.0% | 202.83 | 0.175 |
| Lymphocytes | 8 | 415 | 1703 | -0.59 [-0.88 - (-0.30)] | <0.001 | 0.118 | 79.1% | 33.54 | 0.986 |
| Monocytes | 3 | 59 | 239 | -0.10 [-0.39- 0.19] | 0.519 | 0 | 0% | 0.58 | 0.180 |
| Neutrophils | 4 | 99 | 334 | 0.94 [0.27-1.61] | 0.006 | 0.384 | 85.6% | 20.8 | 0.409 |
| Potassium | 4 | 304 | 1437 | -0.21 [-0.40 - (-0.02)] | 0.034 | 0.015 | 41.2% | 5.1 | 0.502 |
| Procalcitonin | 4 | 194 | 566 | 0.72 [0.06-1.38] | 0.032 | 0.410 | 92.0% | 37.55 | 0.848 |
| Sodium | 4 | 304 | 1437 | -0.26 [-0.67-0.15] | 0.201 | 0.137 | 86.3% | 21.97 | 0.533 |
| Thrombocytes | 7 | 357 | 1621 | -0.57 [-0.68-(-0.45)] | <0.001 | 0 | 0.0% | 3.47 | 0.127 |
| **Others** |  |  |  |  |  |  |  |  |  |
| Time since onset of symptoms to admission | 5 | 236 | 789 | SMD: 0.14 [-0.12-0.41] | 0.291 | 0.056 | 64.8% | 11.36 | 0.465 |
| **Non-survivors (cases) vs survivors (controls)** |  |  |  |  |  |  |  |  |  |
| **Demographics** |  |  |  |  |  |  |  |  |  |
| Sex: male | 7 | 236/340 | 326/617 | 1.32 [1.13-1.54] | 0.005 | 0.002 | 21.8% | 7.67 | 0.70 |
| Sex: female | 7 | 104/340 | 291/617 | 0.65 [0.53-0.83] | 0.005 | 0 | 1.6% | 6.10 | 0.54 |
| Age | 7 | 340 | 617 | SMD: 1.25 [0.78-1.72] | <0.001 | 0.294 | 85.7% | 41.97 | 0.012 |
| **Comorbidities** |  |  |  |  |  |  |  |  |  |
| Any comorbidity | 6 | 207/308 | 234/597 | 1.69 [1.48-1.94] | <0.001 | 0 | 1 | 2.91 | 0.115 |
| Hypertension | 5 | 125/287 | 90/435 | 2.09 [1.65-2.64] | 0.001 | <0.001 | 0% | 2.08 | 0.545 |
| Diabetes mellitus | 5 | 71/318 | 53/451 | 1.88 [1.26-2.81] | 0.012 | <0.001 | 0% | 2.88 | 0.141 |
| Any heart condition | 5 | 48/318 | 15/451 | 3.95 [1.03-15.20] | 0.047 | .477 | 45.5% | 7.35 | 0.666 |
| Cerebrovascular disease | 3 | 12/155 | 0/198 | 36.88 [8.50-160.04] | 0.009 | 0 | 0% | 0.07 | 0.305 |
| Any lung disease | 4 | 39/308 | 14/434 | 3.03 [0.61-15.04] | 0.115 | .429 | 49.8% | 5.97 | 0.811 |
| Carcinoma | 5 | 12/318 | 8/451 | 2.26 [0.67-7.61] | 0.136 | 0 | 0% | 2.84 | 0.02 |
| Current smoker | 4 | 13/200 | 13/322 | 2.02 [0.61-6.72] | 0.160 | <0.001 | 0% | 2.65 | 0.136 |
| **Symptoms and signs** |  |  |  |  |  |  |  |  |  |
| Fever | 6 | 288/319 | 407/455 | 1.00 [0.95-1.05] | 0.974 | 0 | 0% | 4.9 | 0.022 |
| Fatigue | 3 | 109/276 | 129/414 | 1.24 [1.14-1.36] | 0.009 | 0 | 0% | 0.09 | 0.991 |
| Myalgia | 5 | 35/210 | 66/339 | 0.97 [0.61-1.55] | 0.895 | 0.026 | 0% | 3.14 | 0.385 |
| Headache | 4 | 19/255 | 29/301 | 0.83 [0.64-1.09] | 0.120 | 0 | 0% | 0.26 | 0.930 |
| Cough | 6 | 196/319 | 196/455 | 1.37 [0.58-3.24] | 0.385 | 0.605 | 92.3% | 64.86 | 0.389 |
| Sputum | 4 | 84/277 | 93/418 | 1.43 [0.65-3.15] | 0.245 | 0.182 | 62.4% | 7.99 | 0.886 |
| Dyspnea | 4 | 178/264 | 85/314 | 2.60 [0.58-11.65] | 0.137 | 0.561 | 86% | 21.49 | 0.611 |
| Diarrhea | 3 | 48/277 | 71/418 | 0.96 [0.38-2.43] | 0.860 | 0.077 | 27.6% | 2.76 | 0.838 |
| **Treatment** |  |  |  |  |  |  |  |  |  |
| Antibiotics | 5 | 280/309 | 395/438 | 1.03 [0.99-1.07] | 0.114 | 0 | 0% | 2.09 | 0.293 |
| Antiviral treatment | 5 | 222/329 | 446/596 | 0.94 [0.79-1.13] | 0.426 | 0.006 | 67.7% | 12.38 | 0.260 |
| Corticosteroids | 4 | 229/308 | 227/434 | 1.29 [0.66-2.54] | 0.321 | 0.136 | 80.6% | 15.44 | 0.873 |
| Immunoglobulin | 4 | 143/308 | 122/434 | 1.88 [0.36-9.69] | 0.309 | 0.979 | 92.5% | 40.23 | 0.213 |
| Oxygen nasal (high flow) | 4 | 154/308 | 139/434 | 2.16 [0.09-50.50] | 0.493 | 3.843 | 98.1% | 158.98 | 0.03 |
| All mechanical ventilation | 5 | 298/319 | 115/455 | 6.05 [1.41-26.05] | 0.026 | 1.126 | 84.5% | 25.75 | 0.686 |
| Non-invasive mech. ventilation | 5 | 181/309 | 45/438 | 5.33 [1.52-18.71] | 0.021 | 0.565 | 66.7% | 12.02 | 0.765 |
| Invasive mech. ventilation | 5 | 89/309 | 5/438 | 14.14 [138-145.09] | 0.034 | 2.080 | 59.7% | 9.92 | 0.181 |
| Renal replacement therapy | 4 | 22/200 | 1/322 | 10.36 [0.98-110.07] | 0.051 | 0.194 | 0% | 1.92 | 0.057 |
| Extracorporeal membrane oxygenation (ECMO) | 5 | 12/309 | 2/438 | 4.39 [1.64-11.78] | 0.014 | 0 | 0% | 1.35 | 0.033 |
| **Imaging features (CT)** |  |  |  |  |  |  |  |  |  |
| Pathological findings | 6 | 562/577 | 325/335 | 0.97 [0.87-1.09] | 0.588 | 0.006 | 75.9% | 20.71 | 0.675 |
| Pneumonia | 3 | 159/168 | 254/302 | 1.07 [0.97-1.17] | 0.089 | <0.001 | 0% | 1.34 | 0.680 |
| **Complications** |  |  |  |  |  |  |  |  |  |
| Acute respiratory distress symptom (ARDS) | 6 | 298/319 | 115/455 | 4.24 [1.30-13.83] | 0.026 | 1.115 | 92.8% | 69.92 | 0.197 |
| Shock | 4 | 98/277 | 0/418 | 242.79 [23.70-2487.07] | 0.005 | 0 | 0% | 0.64 | 0.300 |
| Acute cardiac injury | 4 | 178/308 | 23/434 | 13.21 [0.70-248.38] | 0.068 | 2.7831 | 81.8% | 16.48 | 0.435 |
| Acute kidney injury | 5 | 88/309 | 5/435 | 20.77 [2.43-177.44] | 0.017 | 2.301 | 67.7% | 12.37 | 0.229 |

| Laboratory parameter | Number of Studies | Number of cases | Number of controls | SMD [95% CI] | p-value | Tau<sup>2</sup> | I<sup>2</sup> | Cochranes Q | Egger's test (p-value)* |
| --- | --- | --- | --- | --- | --- | --- | --- | --- | --- |
| Albumin | 2 | 110 | 120 | -1.14 [-1.41 - (-0.85)] | <0.001 | 0 | 0% | 0 | n.a. |
| Alanine aminotransferase (ALT) | 3 | 223 | 281 | 0.45 [0.08 - 0.82] | 0.016 | 0.056 | 62.7% | 6.37 | 0.984 |
| Aspartate transaminase (AST) | 2 | 114 | 165 | 0.17 [-0.07 – 0.41] | 0.168 | 0 | 0% | 0.76 | n.a. |
| Creatinine | 4 | 200 | 322 | 2.24 [-0.56 – 5.03] | 0.117 | 7.719 | 98.8% | 244.97 | 0.460 |
| C-reactive protein (CRP) | 2 | 114 | 165 | 0 [-0.24 – 0.24] | 1.0 | 0 | 0% | 0 | n.a. |
| D-Dimer | 4 | 174 | 274 | 1.54 [-0.17 – 3.25] | 0.077 | 2.370 | 96.8% | 94.99 | 0.672 |
| Hemoglobin | 3 | 142 | 140 | -0.08 [-0.32 – 0.16] | 0.504 | 0 | 0% | 0.61 | 0.610 |
| Lactate dehydrogenase (LDH) | 2 | 110 | 120 | 1.61 [1.31 – 1.91] | <0.001 | 0 | 0% | 0.3 | n.a. |
| Leucocytes | 4 | 277 | 418 | 2.21 [0.61 – 3.64] | 0.006 | 1.989 | 97.9% | 144.57 | 0.421 |
| Lymphocytes | 4 | 255 | 301 | -0.92 [-1.3 – (-0.55)] | <0.001 | 0.079 | 64.6% | 8.47 |  |
| Neutrophils | 2 | 55 | 141 | 3.6 [3.12 – 4.08] | <0.001 | 0 | 0% | 0.17 | n.a. |
| Potassium | 2 | 55 | 141 | 0.41 [0.1 – 0.77] | 0.01 | 0 | 0% | 0.01 | n.a. |
| Thrombocytes | 4 | 196 | 277 | 0.9 [-2.09 – 3.88] | 0.556 | 8.916 | 99% | 309.32 | 0.487 |
| Partial thromboplastin time (PTT) | 5 | 206 | 294 | 7.99 [4.64 – 11.34] | <0.01 | 13.245 | 98.9% | 370.17 | 0.194 |
| Activated partial thromboplastin time (APTT) | 3 | 65 | 158 | 21.73 [4.34 – 39.13] | 0.014 | 231.933 | 99.5% | 363.82 | 0.386 |
| Interleukin 6 (IL-6) | 2 | 110 | 120 | 1.21 [0.93 – 1.5] | <0.001 | 0 | 0% | 0.44 | n.a. |
| **Others** |  |  |  |  |  |  |  |  |  |
| Time since onset of symptoms to admission | 3 | 195 | 273 | 0.47 [-0.09 – 1.02] | 0.098 | 0.201 | 85.8% | 14.05 | 0.797 |

\*Egger's test cannot be performed with less than three studies. Abbreviation: SMD: Standardize mean difference (negative number indicate lower values in cases, positive number indicate higher number in cases)

**Figure 5.**
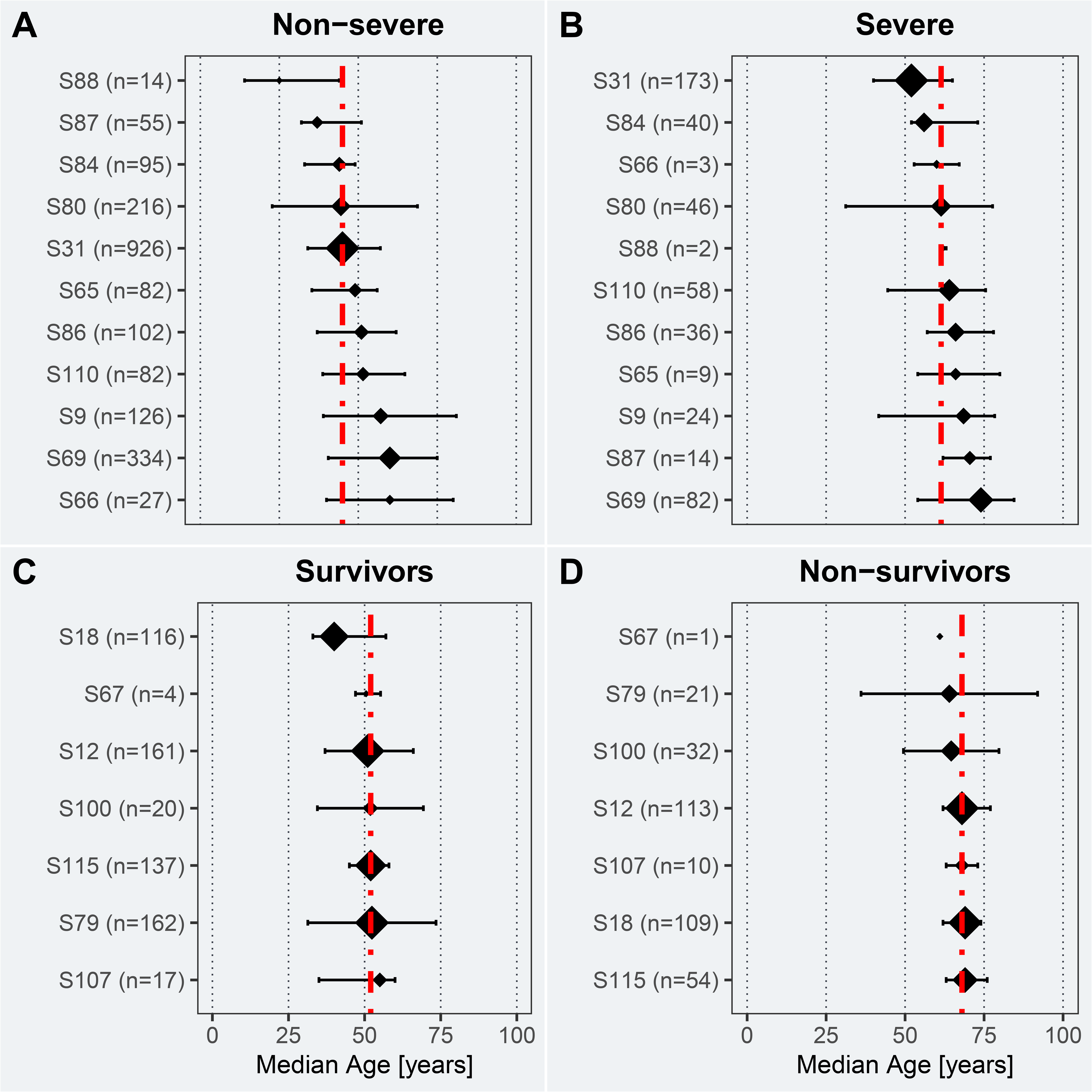
Age of non-severe (A), severe (B), survivor (C), and non-survivor (D) COVID-19 patients included in eligible studies. The median age and Interquartile ranges (IQR) are represented by the midpoints and error bars, respectively. The studies have been sorted by patients’ median age in years. The size of the midpoint indicates the study sample size. The red line indicates the pooled median age of the respective cohort. The key to the study identifier can be found in Table 1.

**Figure 6.**
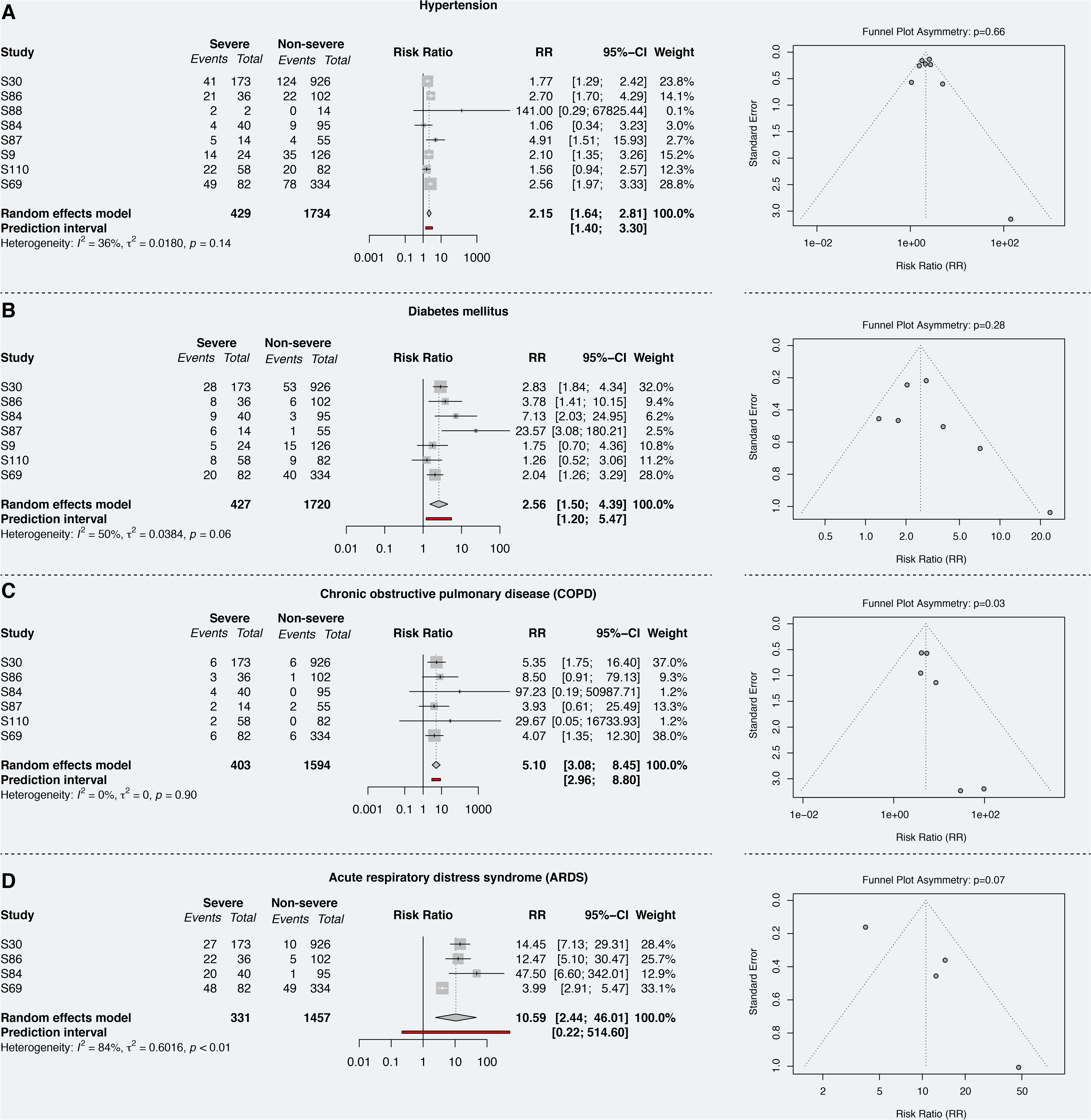
Relative risks of comorbidities (i.e., hypertension, diabetes mellitus, and COPD) and complications(i.e., ARDS) in patients with a severe COVID-19 disease progression. Funnel plots indicate the potential of publication bias. The key to the study identifier can be found in Table 1.

### Survivor vs. non-survivors

Seven studies (957 patients) provided disaggregated data for COVID-19 survivors (617 patients, 64.5%) and non-survivors (340, 35.5%). No differences regarding sex were found in the survivor group (t = 0.258, df = 11.879, p-value = 0.801; male: 326 patients [52.8%] and female: 291 patients [47.2%]), but a significantly higher proportion of male patients were amongst the deceased cohort (t = 4.30, df = 12, p-value = 0.001; male: 236 patients [69.4%] and female: 104 patients [30.6%]) (Supplementary Figure 10). In terms of age, COVID-19 patients that survived were significantly younger (median age in years = 52.0 [35.0–66.0]) than non-survivors (68.0 [62.0–76.0], Figure 5). The meta-analysis yielded older age(SMD: 1.25 [0.78-1.72]; p < 0.001), being male (RR = 1.32 [1.13–1.54], p = 0.005), pre-existing comorbidities (RR = 1.69 [1.48–1.94]; p < 0.001) as potential risk factors of in-hospital mortality. Pre-existing cerebrovascular diseases (RR = 36.88 [8.50–160.04]; p = 0.009), heart conditions (RR = 3.95 [1.03–15.20], p = 0.047, Figure 7A), and hypertension (RR = 2.09 [1.65–2.64]; p = 0.001) were found to be associated with the highest risks of mortality. Clinical signs and symptoms as well as imaging features were comparable between survivors and non-survivors. In terms of treatments, non-survivors were more frequently mechanically ventilated than survivors (RR = 6.05 [1.41–26.05]; p = 0.026, Figure 7B) and more commonly received extracorporeal membrane oxygenation (RR = 4.39 [1.64–11.78], p = 0.014). Non-survivors had higher risks of complications, particularly acute kidney injury (RR = 20.77 [2.43–177.44], p = 0.017; Figure 7C) and ARDS (RR = 4.24 [1.30–13.83], p = 0.026, Figure 7D). Low levels of albumin (SMD = –1.13 [−1.41 – (−0.85)]; p < 0.001) and lymphocytes (SMD = –0.92 [−1.3 – (−0.55)]; p < 0.001) as well as elevated level of interleukin 6 (SMD = 1.21 [0.93 – 1.5]; p < 0.001), leucocytes (SMD = 2.21 [0.61 – 3.64]; p = 0.06), and prolonged prothrombin time (SMD = 7.99 [4.64 – 11.34]; p < 0.01) were associated with death (Table 5). Publication bias, measured by means of the Egger’s test, was only evident in five analyses.

**Figure 7.**
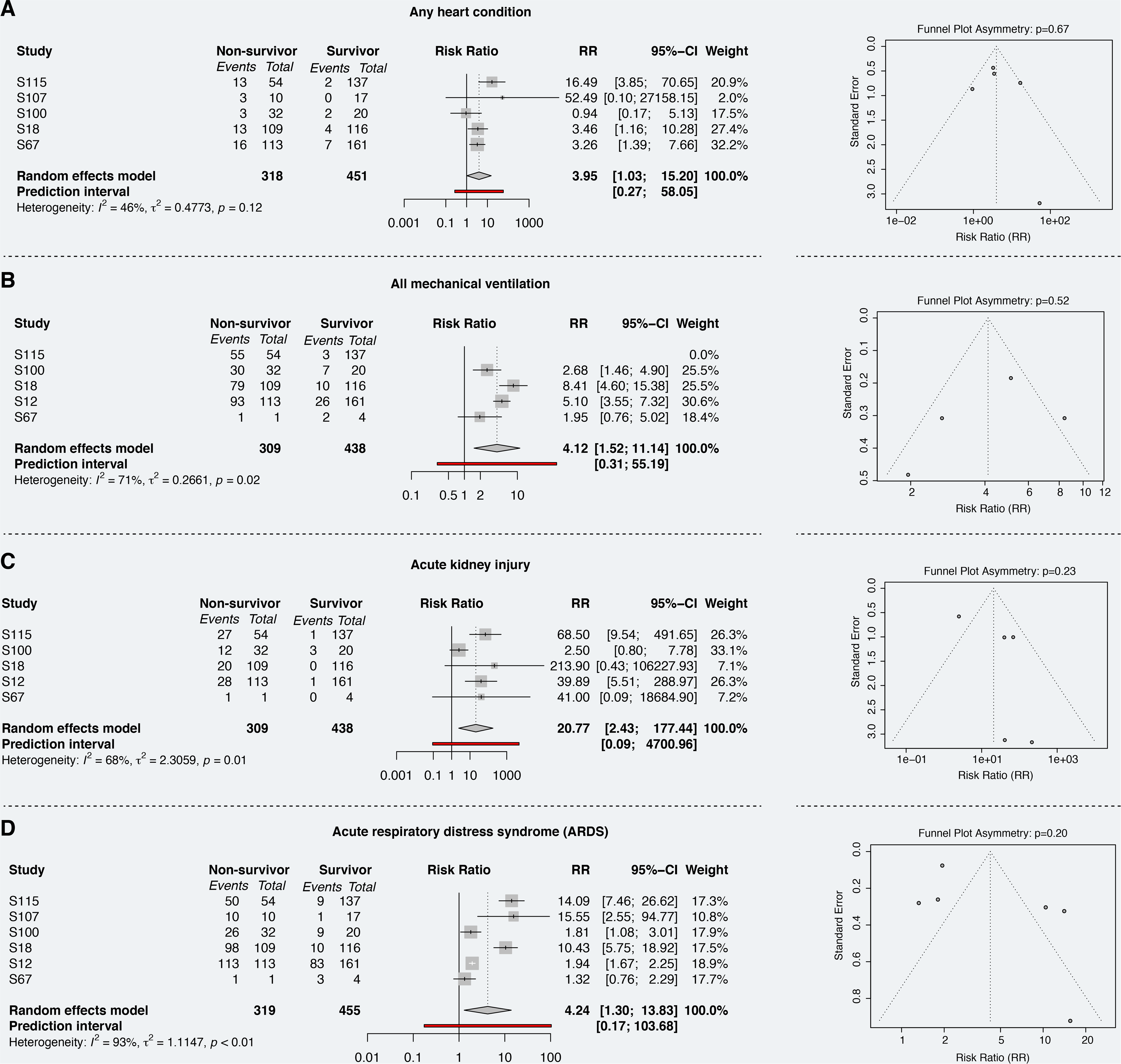
Relative risks of comorbidity (i.e., any heart condition), treatment (i.e., mechanical ventilation), and complications (i.e., acute kidney injury and ARDS) in survivors and non-survivors. Funnel plots indicate the potential of publication bias. The key to the study identifier can be found in Table 1.

## 4. Discussion

As of May 1^st^, 2020, more than 3.3 million confirmed cases of COVID-19 and more than 230’000 deaths attributable to the disease, have been reported worldwide^166,167^. In-depth knowledge of clinical, laboratory, and imaging factors that are associated with the disease progression and outcome is critical to inform clinical decision making and pandemic preparedness initiatives. An ever-growing number of research studies have been performed, but thus far the meta-analytical evidence is sparse. To address this paucity, we conducted a systematic review and meta-analysis of 148 studies involving over 12’000 patients providing an unprecedently comprehensive overview of comorbidities, clinical signs and symptoms, laboratory parameters, CT imaging features, treatment, outcomes, and complications in adult, pregnant, and pediatric/neonatal COVID-19 patients. Approximately eight percent of the patients were reported to be asymptomatic, while over seven percent died from complications associated with COVID-19. Recent analysis suggests that up to 75% of the coronavirus infections caused no illness^168–170^. Presumably, the virus has been circulating for longer than generally believed and large swathes of the population have already been exposed. Although our fatality rate lies within previous estimates^171,172^, it is important to mention that only a limited number of studies reported on the outcome of COVID-19 (i.e., death, survival, recovery) and thus, caution has to be exercised when interpreting this number. Through our meta-analysis, we revealed several important risk factors that are associated with severe disease progression and mortality. Among these risk factors were two demographic factors, namely older age and being male. Well-studied consequences of ageing are the decline in the immune function (e.g., T-cell and B-cell function) and excess production of type 2 cytokines^173,174^. These age-dependent changes in the immune response are suspected to cause deficiency in control of viral replication and more prolonged proinflammatory responses, potentially leading to poor outcome^175^. Corroborative evidence stems from preclinical studies that found an age-dependent host innate responses to virus infection in non-human primates inoculated with SARS-CoV-1^176^. Confirming previous findings^177,178^, sex-specific differences in mortality and vulnerability to the disease were evident in the current study. Specifically, men were disproportionately affected by an infection with SARS-CoV-2 (i.e., proportion of men presented with COVID-19 was larger compared to women) and the in-hospital mortality amongst male patients was significantly higher compared to female patients. Emerging evidence pinpoints towards differences in the immune system^140^, genetic polymorphism^179^, life style factors including smoking^180^, personal hygiene habits^181^, pre-existing comorbidities^182,183^, and expression of angiotensin-converting enzyme 2 (ACE2)^184,185^ as potential explanations for the increased vulnerability in men. This sex difference in vulnerability has also been observed for SARS and MERS^186^, two previously emerging coronavirus diseases. The lack of sex-disaggregated data in the reviewed studies made it impossible to further explore these potential explanations for the discrepant findings in men and women. Overall, the preexisting comorbidities, namely hypertension, diabetes mellitus, and any heart condition, were found to be linked with both, more severe diseases status and increased in-hospital mortality. Smoking, by contrast, was not associated with disease severity or mortality. However, the low number of studies reporting smoking status (13/ 148) cautions against early assumptions. Clinical signs and symptoms were comparable between patients with non-severe and severe COVID-19 as well as survivors and non-survivors. Fever, cough, and myalgia were amongst the most frequent reported symptoms across all groups. Similarly, the present study revealed no differences in the CT imaging features. The majority of the COVID-19 patients presented with pneumonia (bilateral or unilateral) and GGO. These pathological findings are a hallmark of any viral pneumonia, and thus it is not surprising that asymptomatic patients had similar distinctive features^187^. In terms of laboratory parameters, elevated levels of interleukin 6, leucocytes, d-dimer, and lactate dehydrogenase as well as hypoalbuminemia and lymphopenia were more commonly seen in patients with severe COVID-19 illness and non-survivors. High levels of d-dimer have a reported association with 28-day mortality in patients with infections or sepsis admitted to the intensive care unit^188^. Systemic pro-inflammatory cytokine responses (e.g., interleukin 6 and other components) contribute to host defense against infections, such as SARS-CoV-2^189–191^. However, exaggerated synthesis of interleukin 6 can lead to an acute, severe systemic inflammatory response syndrome (SIRS) known as ‘cytokine storm’^192^. In addition to SIRS, hypoalbuminemia and lymphopenia were previously shown to be associated with increased odds of severe infection and infection-related death^193–195^. Complications were very common amongst patients with severe COVID-19 disease (over 50%) and non-survivors (more than two thirds). Acute cardiac injury, ARDS, and acute kidney injury were strongly linked to the outcome. Widely used treatments for COVID-19 and associated complications comprised antibiotics, antivirals, and oxygen therapy. Patients with severe COVID-19 disease required more often mechanical ventilation and renal replacement therapy compared to those with non-severe COVID-19. Moreover, corticosteroids have been commonly administered to hospitalized patients with severe illness, although their benefit is highly disputed. Evidence from MERS or influenza suggests that patients who were given corticosteroids had prolonged viral replication, receive mechanical ventilation, and have higher mortality^196–199^. Administration of antibiotics and antivirals was independent of disease-severity.

Pregnant women as well as pediatric and neonatal patients may be less vulnerable to complications of COVID-19. Comorbidities were almost non-existent in these patient cohorts. Clinical signs and symptoms, laboratory parameters, imaging features, and treatments were comparable to the adult (non-pregnant) cohort. While there was a considerable proportion of children and neonates with SARS-CoV-2 infections reported, most of these patients did not need hospitalization and recovered quite well. With the exception of a 10-month old neonate, no children were amongst the deaths reported. All pregnant women included in our study survived COVID-19 and associated complications.

### 4.1. Limitations of review

A limitation of the current review was that literature search was limited to articles listed in EMBASE, PubMed/ Medline, Scopus, Web of Science, or identified by hand searches. Considering the pace at which the research in this area is moving forward, it is likely that the findings of the publications described in this paper will be quickly complemented by further research. The literature search also excluded grey literature (e.g., preprints, reports, conference proceedings), the importance of which to this topic is unknown, and thus might have introduced another source of search bias. There is also a probability of publication bias, as well as potential for a search bias. Publication bias is likely to result in studies with more positive results being preferentially submitted and accepted for publication. Moreover, geographical bias cannot be rule out as the majority of the studies (129/ 148) were conducted in China. While symptoms might be quite comparable across countries, comorbidities, treatments, and outcome potentially depends on the country (and its healthcare system). There is also a considerable risk for a reporting bias towards comorbidities, clinical signs and symptoms, laboratory parameters, imaging features, treatment, outcome, and complications that are present. Specifically, only a minority of studies reported a zero when this information was assessed, but absent in patients. Lack of data on absent clinical signs and symptoms might lead to distorted estimates of proportion. The meta-analysis of severity and mortality could only be performed with a small number of studies as the minority of the 148 provided data separately for different disease severity groups (e.g., non-severe, severe, survivors, non-survivors). This needs to be considered when interpreting the results, including the publication bias as the Egger’s test may lack the statistical power to detect bias when the number of studies is small (i.e., < 10). Lastly, the criteria to classify patients in severe and non-severe COVID-19 disease cohorts varied between studies leading to additional heterogeneity between studies. By virtue of low number of studies available, we could not assess this heterogeneity nor adjust for it.

### 4.2. Conclusion and future directions

In conclusion, this unprecedentedly comprehensive systematic review and meta-analysis of the literature published during the first 120 days of the COVID-19 pandemic yields important information regarding the comorbidities, clinical signs and symptoms, laboratory parameters, imaging features, treatment, outcome, and complications. Male sex, older age, and pre-existing comorbidities are major risk factors for in-hospital mortality and complications. This study revealed a fatality rate of 7.7% and found that approximately 8% of the patients were reportedly asymptomatic. Based on recent reports, the latter number is likely 6- to 10-fold higher as only a few asymptomatic patients are captured by the health care system as they do not seek medical attention due to the lack of symptoms^168^ or are not hospitalized and thus, included in studies. Unnoticed asymptomatic cases of COVID-19 are likely a major source of ongoing transmission. Children and neonates appear to be the least vulnerable cohort. Forthcoming studies are needed that provide sex-disaggregated data to better characterize risk factors that affect both sexes or are specific to men or women, respectively.

## Data Availability

The code used for the analysis and to create figures and tables is provided in our GitHub repository (https://github.com/jutzca/Corona-Virus-Meta-Analysis-2020).

## Authors’ contribution

### Catherine Jutzeler

Substantial contributions to the conception and design of the study; acquisition, analysis, and interpretation of data, drafting the manuscript, final approval of version to be published.

### Lucie Bourguignon

Substantial contributions to acquisition, analysis, and interpretation of data, drafting the manuscript, final approval of version to be published.

### Caroline Weis

Acquisition and interpretation of data, revising the manuscript critically for important intellectual content, final approval of version to be published.

### Bastian Rieck

Acquisition and interpretation of data, revising the manuscript critically for important intellectual content, final approval of version to be published.

### Bobo Tong

Acquisition and interpretation of data, revising the manuscript critically for important intellectual content, final approval of version to be published.

### Cyrus Wong

Acquisition and interpretation of data, revising the manuscript critically for important intellectual content, final approval of version to be published.

### Hans Pargger

Substantial contributions to the interpretation of data, revising the manuscript critically for important intellectual content, final approval of version to be published.

### Sarah Tschudin-Sutter

Substantial contributions to the interpretation of data, revising the manuscript critically for important intellectual content, final approval of version to be published.

### Adrian Egli

Substantial contributions to the interpretation of data, revising the manuscript critically for important intellectual content, final approval of version to be published.

### Karsten Borgwardt

Substantial contributions to the interpretation of data, revising the manuscript critically for important intellectual content, final approval of version to be published.

### Matthias Walter

Substantial contributions to the conception and design of the study; acquisition, analysis, and interpretation of data, drafting the manuscript, final approval of version to be published.

## 4.3. Conflict of interest

The authors do not report any (financial or otherwise) conflict of interest.

## 4.4. Acknowledgement

This study was supported by the Alfried Krupp Prize for Young University Teachers of the Alfried Krupp von Bohlen und Halbach-Stiftung (Borgwardt), and the Swiss National Science Foundation (Ambizione Grant [PZ00P3_186101, Jutzeler], SNSF Starting Grant [155913, Borgwardt}). The funders had no role in study design, data collection and analysis, decision to publish, or preparation of the manuscript.

## Notes

### Competing Interest Statement

The authors have declared no competing interest.

